# Sonic hedgehog inhibitor suppresses carcinoma associated fibroblasts to prime Gemcitabine/Nab-Paclitaxel and anti-CTLA4 immunotherapy as sequential first-line combination therapy in a Phase 1b/2 study in PDAC: NUMANTIA trial

**DOI:** 10.64898/2025.12.14.25342225

**Authors:** Valerie S. Kalluri, Bruno Bockorny, Evelio Perea Borobio, Teresa Macarulla, Roberto Pazo Cid, Laura Medina, Aitziber Gil-Negrete, Fernando Rivera, Vanesa Varela, Alejandro Martin-Munoz, Yanira Ruiz-Heredia, Bingrui Li, Patience Kelly, Barbara Moreno Diaz, Shreyasee V Kumbar, Hikaru Sugimoto, Raghu Kalluri, Manuel Hidalgo

**Author notes:** **Corresponding Authors:** Manuel Hidalgo, MD, PhD; NYU Grossman School of Medicine, New York, NY;, Raghu Kalluri, MD, PhD; University of Texas MD Anderson Cancer Center, Houston, TX.

## Abstract

Metastatic pancreatic ductal adenocarcinoma (PDAC) remains deadly, with minimal improvement in prognosis over the past 20 years despite expansion of our chemotherapeutic arsenal. The complex tumor microenvironment (TME) of PDAC in the advanced stage, which often accompany clinical diagnosis, likely contributes to the limited efficacy of current standard of care chemotherapy. Informed by mechanistic preclinical studies, we evaluated the impact of inhibition of Hedgehog (Hh) signaling to prime PDAC TME and leverage anti-tumor efficacy of Gemcitabine plus nab-Paclitaxel (GnP) together with anti-CTLA-4 immune check point inhibitor (ICI, zalifrelimab). Hh inhibition using NLM-001 (an oral small molecule inhibitor of Smo) aimed to polarize the PDAC TME, including cancer associated fibroblasts (CAFs) and intratumoral immune profile, and to foster an immunosuppressive milieu that engages ICI. In this Phase 1b/2, open label, single arm study, patients with metastatic PDAC received standard GnP every 28-day cycles. In addition, NLM-001 was given at 800 mg daily on days -4 to -1 and days 10 to 13 of the GnP cycles 1 to 3, 6 to 8, 11 to 13 onwards (3 cycles on, followed by 2 rest cycles). Anti CTLA-4 inhibitor, zalifrelimab, was administered at 1mg/kg on day 15 of cycle 1 and every 6 weeks thereafter. The primary end point was to assess efficacy by objective response rate (ORR) as per RECIST v1.1. Treatment was overall well tolerated in the 28 patients enrolled. Most frequent grade 3-4 adverse events (AEs) were neutropenia (46.4%), asthenia (21.4%), and neurotoxicity (14.3%). No patient discontinued treatment due to toxicity. ORR was 50% [95% CI, 29.1-70.9] and disease control rate was 95.5% [95% CI, 86.8 - 100.0]. Median progression-free survival (PFS) was 7.3 months 95.5% [95% CI, 5.564 – 9.041] and median overall survival (OS) was 11.5 months [95% CI, 10.23 –12.73]; 1-year PFS was 18.2% [95% CI, 2.1 – 34.3] and 1-year OS was 50% [95% CI, 29.0 – 710]. Patients who achieved ctDNA clearance at cycle 4 had a significant better PFS (10.7 vs 6.0 months; p<0.0001). Paired biopsies immunolabeling and spatial transcriptomic analyses showed polarization of the TME, with increased CD4^+^ and CD8^+^ T cells infiltration, down trending Tregs, and decreased αSMA/FAP ratio. Hedgehog inhibitor NLM-001 in combination with gemcitabine/nab-paclitaxel and zalifrelimab was safe and well tolerated and showed encouraging objective responses in the first line treatment of advanced PDAC.

**Clinical Trial Registration:** EudraCT: 2020-004932-52; NCT04827953.

## Introduction

Pancreatic ductal adenocarcinoma (PDAC) is an aggressive malignancy with treatment options still largely limited to systemic chemotherapies^1,2^. Minimal impact has been realized to improve survival outcome despite our growing understanding of the genetic and stromal component of PDAC^2–4^. The PDAC tumor microenvironment (TME) is now recognized as a dynamic component of cancer progression and therapeutic resistance, with both promoting and restraining functions^3,5–10^. The functional heterogeneity of cancer associated fibroblasts (CAFs) and their role in shaping the tumor immune microenvironment represents a significant lever in realizing therapeutic benefit of immune modulating therapies^3,10^. Efforts to target CAFs contributed to our understanding of the PDAC TME, and associated Hedgehog (Hh) signaling as a possible regulator of CAFs function in PDAC^11^. Hedgehog ligands bind to the receptor protein Patched (Ptch) and relieve its suppression of Smoothened (Smo), leading to downstream activation of glioma-associated oncogene homolog 1 (Gli1) transcriptional effects, sustaining proliferation and suppressing cell death^12^. Hh signaling had been implicated in reshaping the immunosuppressive microenvironment of PDAC, stimulating CAFs-mediated extracellular matrix deposition^13–18^.

Preclinical studies pointed to a transient benefit of Hh inhibition in PDAC in mice by enhancing vascular perfusion of tumors and gemcitabine efficacy, and this was associated with suppression of αSMA^+^ CAFs^19^. However, lasting benefit was not achieved due to its unintended impact on the TME^20^. Inhibition of Hh signaling failed in patients with PDAC, stopping a Phase 2 trial of Hh inhibitor IPI-926 combined with gemcitabine due to worse outcomes in the IPI-926 arm compared to placebo^21^. In genetically engineered mice (GEM) with spontaneous PDAC, Hh pathway inhibition was observed to influence CAFs and was associated with increased Treg and decreased cytotoxic T cells, generating an immunosuppressive TME^18^. Genetic targeting of αSMA^+^ CAFs in mice similarly led to more poorly differentiated tumors and generated an immunosuppressive TME with increased Tregs^22^. Conditional deletion of sonic Hh in PDAC GEM mimicked the histopathology of αSMA^+^ CAFs depleted tumors and showed reduced αSMA^+^ CAFs content^20^. Collectively these preclinical studies offered insights into the early clinical failure of Hh inhibition in PDAC: although the targeting of PDAC TME with Hh inhibitor enabled transient perfusion of chemotherapy, the net effect was the generation of an immunosuppressive, tumor promoting milieu with decreased survival. These studies however also informed on Hh inhibition in creating an opportunistic vulnerability to leverage immune checkpoint therapy. In PDAC GEM, treatment with anti-CTLA4 ICI in the context of αSMA^+^ CAFs depletion demonstrated increased survival benefit^22^. The remodeling of PDAC stroma thus appears to create novel opportunity for ICI efficacy, which had showed otherwise limited impact in patients with PDAC^23^.

NLM-001 (previously known as TAK-441) is an investigational small molecule that is administered orally and directly inhibits Smo^24^. NLM-001, studied in a phase I trial on patients with advanced solid tumors including pancreatic cancer, was found to be safe and well tolerated, and was noted to inhibit Gli1 gene expression in skin biopsies^25^. Informed by mechanistic preclinical studies described above that implicate a context dependent impact of Hh targeting on PDAC CAFs, we designed a clinical trial in which priming of the TME with NLM-001 phasic dosing was utilized to favor response to chemotherapy and ICI. Here, we report the results of the phase 1b/2 with hedgehog inhibitor NLM-001 in combination with gemcitabine/nab-paclitaxel, and CTLA-4 inhibitor zalifrelimab in the first line for metastatic pancreatic cancer.

## Results

### Trial design for Hh inhibition priming with ICI and chemotherapy combination

Inhibition of Hh signaling with cyclopamine^26^ in PDAC GEM recapitulated the more poorly differentiated histopathology noted with αSMA^+^ CAFs depletion^22^, and concurrent treatment with anti-CTLA4 and anti-PD1 ICI showed improved histopathology compared to controls (**Extended Data Fig. 1**), supporting that the priming effect of Hh inhibition on PDAC TME lends itself to ICI response. Targeting of αSMA^+^ CAFs, by direct genetic means^22^ or via conditional Hh signaling targeting in cancer cells^20^, informed on opportunistic TME remodeling to sensitize PDAC to ICI. Taken together, the trial design set forth combined priming of the TME with Hh inhibitor, followed by gemcitabine/nab-paclitaxel (GnP) and anti-CTLA4 (zalifrelimab) ICI. A total of 40 patients with untreated advanced PDAC were consented for the study. Twelve subjects failed screening, 11 due to not meeting inclusion/exclusion criteria and 1 due to consent withdrawal. Twenty-eight subjects were enrolled and received treatment and were considered the intention to treat (ITT) population (**Fig. 1A**). The mean age of the population was 59.2 years (IQR range 53.0 – 67.5), and 35.7% were female (**Table 1**). ECOG performance status was 0 in 53.6% and 1 in 46.4%, and all patients had histologically confirmed adenocarcinoma (**Table 1**). Twenty-seven patients had *de novo* stage IV disease, and one had stage III at diagnosis. All but one patient had untreated metastatic disease at enrollment with 89.3% having liver metastasis.

**Figure 1.**
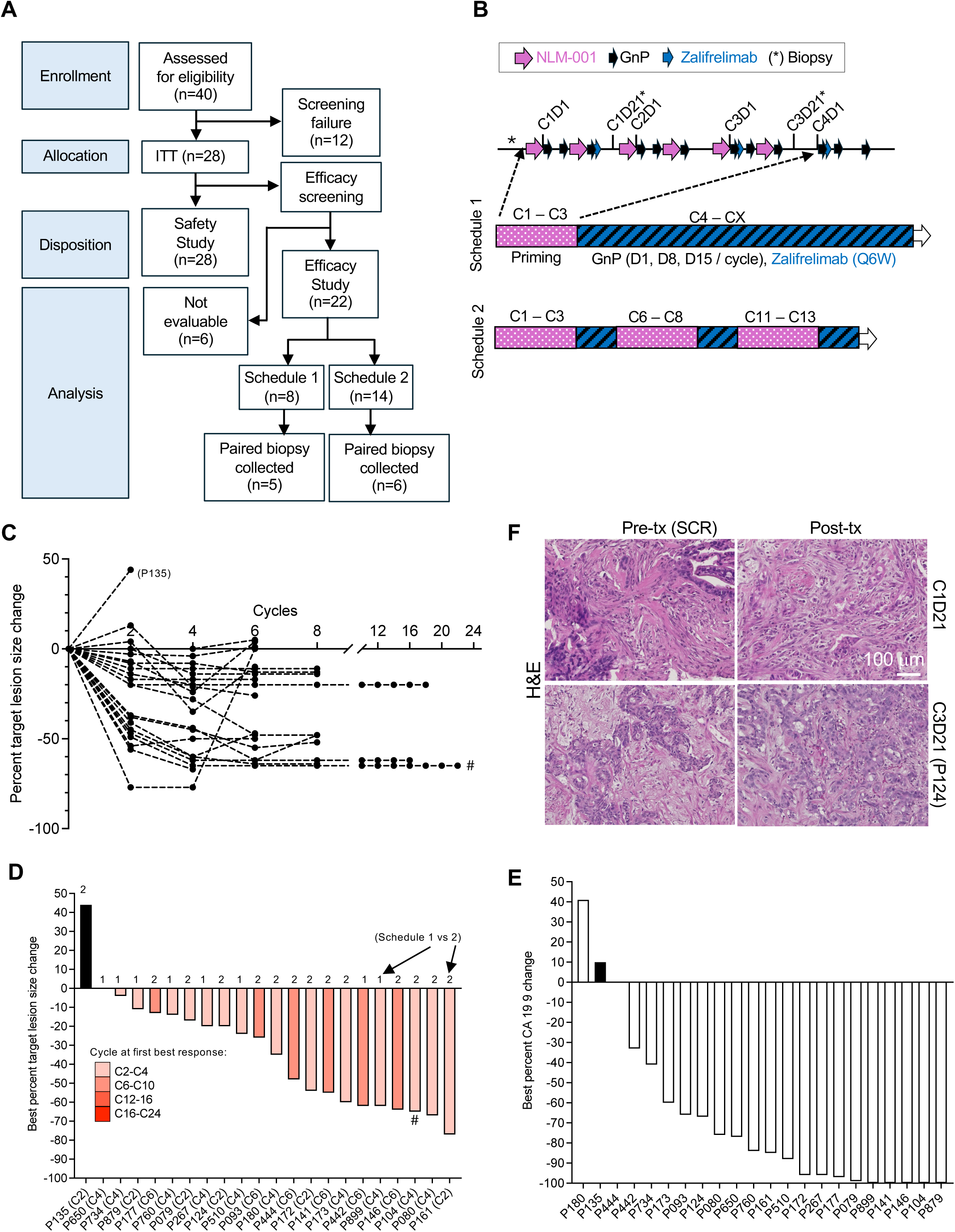
NUMANTIA trial design and endpoint. **A.** CONSORT diagram. **B**. Schematic representation of dosing, with repeated NLM-001 priming cycles in schedule 2 (C1-C3, C6-C8, C11-C13, C16-C18 and so on) versus a single priming in schedule 1 (C1-C3). **C**. Percent change in target lesions ascertained overtime for each patient on study. Subject P135 is denoted as the single patient with early progression. #: P104, no progression on study. **D**. Bar graph depicting best percent target lesion size change for each patient. The numbers ‘1’ and ‘2’ refers to schedule 1 vs 2 allocation. Color scheme indicates treatment cycle range at which best response was achieved. #: P104, no progression on study. **E**. Bar graph depicting best percent change in CA 19-9 for each patient listed. **F**. Representative H&E micrographs of pre-treatment (pre-tx, SCR = screening) and post-treatment (post-tx). Top row depicts paired biopsies captured at C1D21; bottom row depicts paired biopsies captured at C3D21 (for one patient, P124).

**Table 1.**
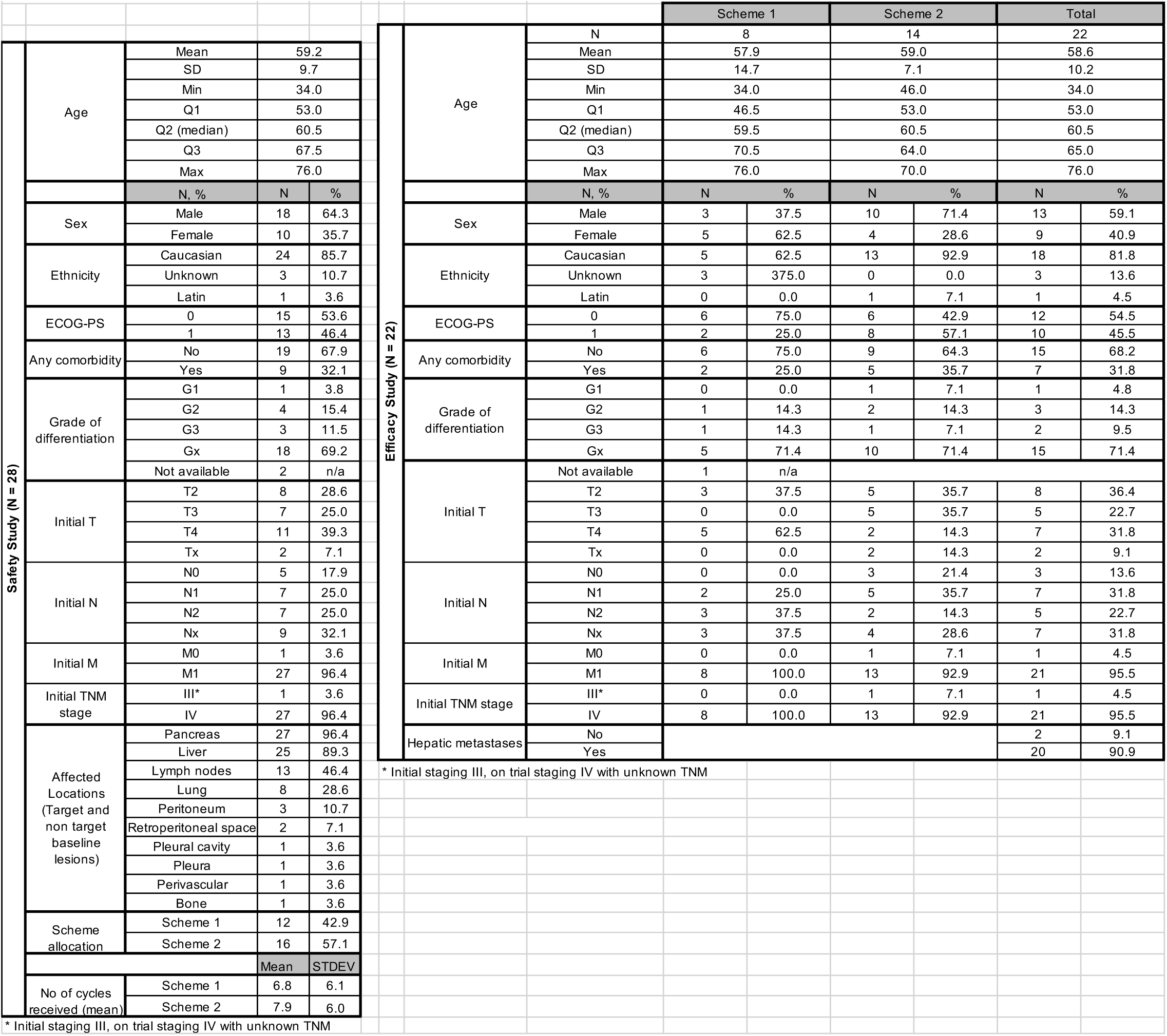
Patient characteristics.

Patients received conventional chemotherapy with gemcitabine (1,000 mg/m^2^ IV) and nab-paclitaxel (125 mg/m^2^ IV) on days 1, 8 and 15 of each 28-day cycle according to standard of care (**Fig. 1B, Extended Data Fig. 2A**). Zalifrelimab was given at a dose of 1mg/kg IV on day 15 of cycle 1 and every 6 weeks, 30 min after administration of chemotherapy (**Fig. 1B, Extended Data Fig. 2A**). NLM-001 was administered at the dose of 800 mg/day orally according to two schedules. The subjects were initially enrolled to receive “Schedule 1” in which NLM-001 was given for 4 days as a priming treatment before chemotherapy days 1 and 15 (that is, on days - 4, - 3, - 2 and - 1 and days 10, 11, 12, and 13), for cycles 1-3 (**Fig. 1B, Extended Data Fig. 2A**). Interim analysis indicating safety and preliminary responses in the first 12 patients enrolled in “Schedule 1” prompted protocol amendment to include additional NLM-001 priming, with subsequent patients to receive treatment according to “Schedule 2”, in which NLM-001 was given for three consecutive cycles followed by two rest cycles (cycles 1-3, 6-8, 11-13, 16-18 and beyond) (**Fig. 1B**, **Table 1, Extended Data Fig. 2A**). Paired biopsies were obtained, when feasible, for assessment of translational endpoints prior to initiation of treatment (screening) and at the completion of Cycle 1 Day 21; except for one sample (P124) captured at Cycle 3 D21. Imaging was performed Q6W (±3 days) from the first treatment dose. (**Fig. 1B**, **Table 1, Extended Data Fig. 2A, Extended Data Table 1**).

### Priming with Hh inhibition with ICI and chemotherapy combination was well tolerated

Safety is reported for all 28 patients who received at least one dose of study drug in the safety analysis set (**Fig. 1A**). All patients received NLM-001, while 27 received gemcitabine and nab-paclitaxel, and 24 received zalifrelimab (**Extended Data Table 2**). All patients experienced at least one treatment-emergent adverse event (TEAE, **Supplementary Table 1**) with the most common being asthenia in 82.1% (grade 3 in 21.4%), neutropenia in 67.9% (grade 3 and 4 in 46.4%), nausea in 64.3%% (grade 3 in 7.1%) and neurotoxicity in 46.4% (grade 3 in 14.3%) (**Supplementary Table 1**). The most common treatment related AE (TRAE) related to NLM-001 (alone or in combination) were asthenia in 40.9% (grade ≥ 3 in 4.5%), nausea in 27.3% (all grade 1-2), anorexia in 18.2% (grade 3 in 4.5%), and neutropenia in 18.2% (grade 3 in 4.5%) (**Table 2**). Specific immune-mediated TRAEs related to zalifrelimab, namely immune-mediated enterocolitis, immune-mediated polyserositis, pneumonitis, or rash were noted in 4.5% of patients (n = 22 patients, **Extended Data Table 3**). Serious AEs (SEAs) were encountered in 36.4% of patients and all resolved with close medical supervision, with or without dose omission or delay (**Supplementary Table 2**). There were no treatment-related deaths in the study.

**Table 2.**
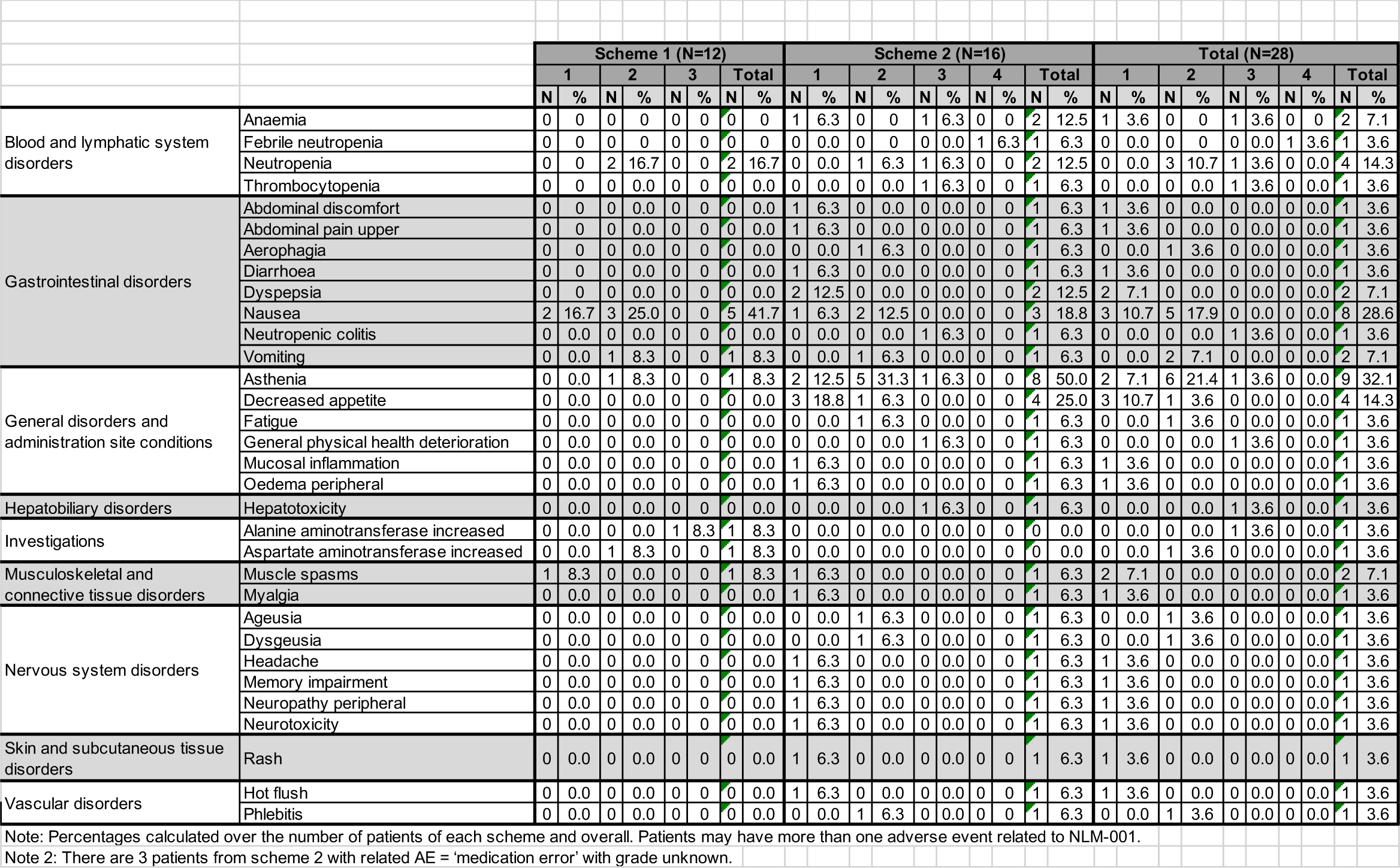
Adverse events related to NLM-001 (alone or in combination with other study drugs) by max grade and scheme.

### Efficacy analyses indicate robust clinical activity in the majority of patients

Efficacy-evaluable set included all treated patients who received at least two complete cycles of study treatment as specified a priori (n = 22). Eight patients received treatment according to schedule 1 and 14 according to schedule 2 (**Fig. 1A**). The median treatment exposure was 35.9 weeks (range, 8.7-106.7). A total of 11 of 22 evaluable patients achieved a response according to RECIST 1.1, with ORR of 50% (95% CI, 29.1 – 70.9) (**Fig. 1C**). Disease control was achieved in 21 patients (95% CI, 86.8 - 100.0) (**Fig. 1C**). One patient showed progressive disease (**Fig. 1C**). Of the 11 patients with partial response, 7 achieved best target lesion reduction within Cycle 2 to Cycle 4 of treatment (**Fig. 1D**). Nine out of 11 patients with partial response followed Schedule 2 dosing vs 2 out of 11 followed Schedule 1 dosing (**Fig. 1D**, **Extended Data Fig. 2B-C**). No significant differences were noted in overall survival (OS) and progression free survival (PFS) in patients following Schedule 1 vs Schedule 2 of treatment (**Extended Data Fig. 2D**). Median Duration of Response (DOR) was 8.3 months (95% CI, 0.3 – 16.3). The median PFS was 7.3 months (95% CI, 5.6 – 9.0) and the 6-month PFS was 77.3% (95% CI, 59.9 –94.7). Median OS was 11.5 months (95% CI, 10.2 – 12.7), and 12-month OS was 50.0% (95% CI, 29.0 – 71.0). One patient did not show progression while on study (#, **Fig. 1C-D**).

Analyses of circulating tumor marker CA19-9 support response to therapy, with 17 out of 20 patients with elevated CA19-9 at baseline (85%) showed a decrease in CA19-9 greater than 50% from baseline, and 70% of patients experienced a reduction greater than 75% (**Fig. 1E**). CA19-9 decrease was noted in patient with minimal response characterized by RECIST 1.1 (**Extended Data Fig. 2E**). Representative H&E of pre-treatment (screening, SCR) and post-treatment (C1D21 and C3D21 for P124) show more differentiated histopathological features (**Fig. 1F**), with stromal remodeling described therein (vide infra).

### Early ctDNA declines are associated with therapeutic response

Twenty-two patients were followed longitudinally for molecular responses evaluated by mutation specific ctDNA quantification (**Fig. 2A**). One or more somatic variants of known association with PDAC were assessed (**Extended Data Table 4**). At completion of Cycle 4, when most patients with response were noted on imaging (**Fig. 2B**), those who achieved ctDNA clearance had a significantly better median (10.7 versus 6.0 months with a favorable AUC ratio >1, p<0.0001) and an improved OS (12.5 versus 8.7 months, AUC ratio >1, p=0.0527) (**Fig. 2C**).

**Figure 2.**
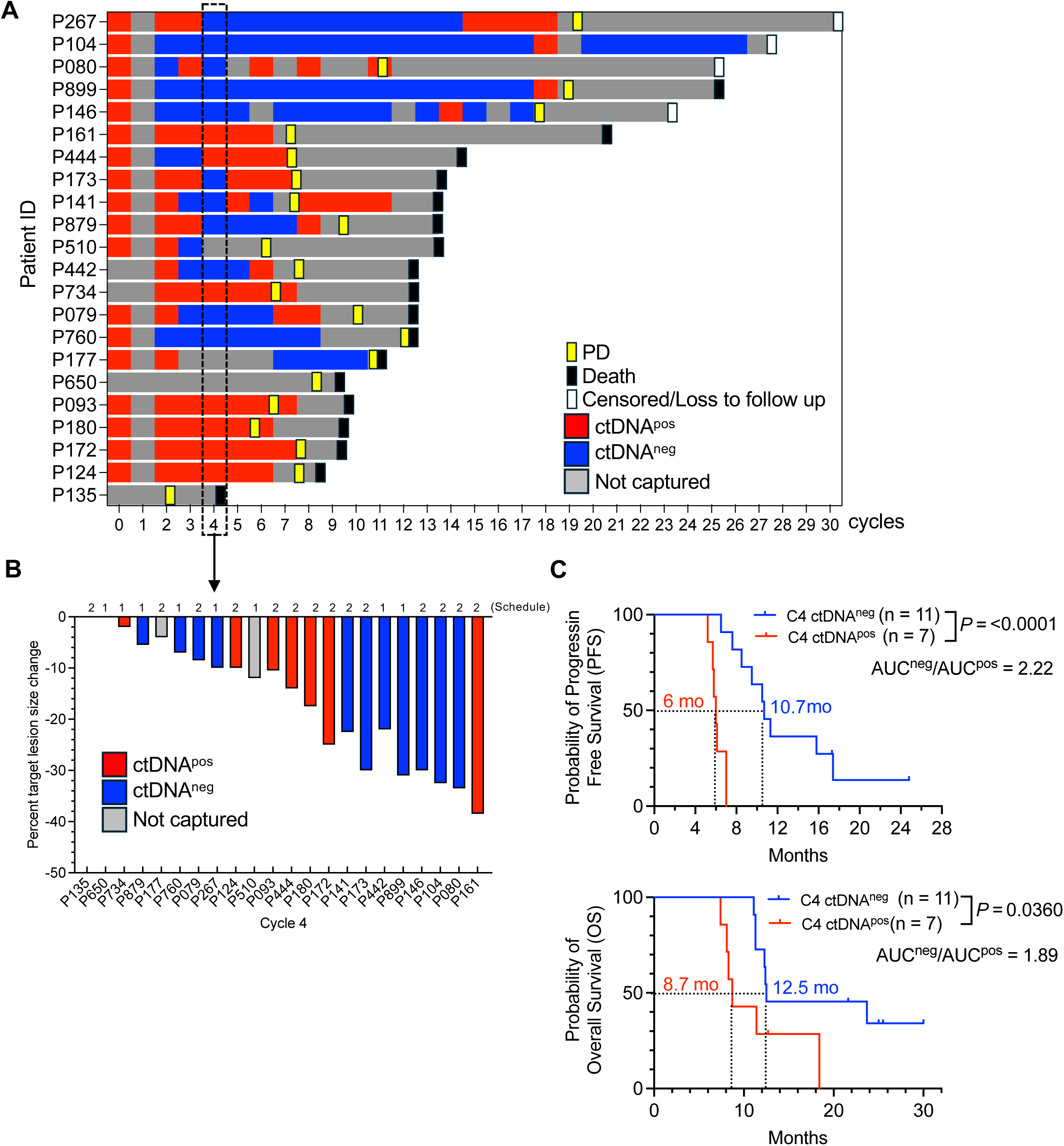
ctDNA for defined PDAC mutations informs PFS and OS. **A**. Schematic representation of ctDNA positive (red) and negative (blue) sampling over time for each patient over time, listed as cycles of treatment. Endpoint (death, progression of disease (PD) or censoring) is defined for each patient. **B**. Bar graph depicting best percent target lesion size change for each patient at cycle 4 of treatment. The numbers ‘1’ and ‘2’ refers to schedule 1 vs 2 allocation. Color scheme indicates ctDNA positive vs negative status at cycle 4. **C.** Progression free survival (PFS) and overall survival (OS) of patients with positive vs negative ctDNA at cycle 4. Gehan-Breslow-Wilcoxon test, *P* values are listed. Mo: months. AUC: area under the curve; AUC ratio is listed.

### Hh priming polarizes the PDAC TME

Evaluation of biopsies taken pre-treatment initiation and at C1D21, 7 days after completion of Hh priming x2, GnP x3, and zalifrelimab x1 (**Fig. 1B, Extended Data 2A**). For P124, the post-treatment initiation biopsy was captured at C3D21 (**Extended Data Table 1**). All but one pair of biopsies were liver metastases, one was from a supraclavicular lymph node (LN) (P267). Moderate increase in collagen abundance, visualized by Sirius red staining, was noted post-treatment (**Fig. 3A**). Polarized light microscopy indicated remodeling of the collagen fibers with a more interrupted/less linear pattern post treatment (**Fig. 3B**). Immunolabeling for αSMA^+^ and FAP^+^ CAFs revealed similar/mild increase in αSMA^+^ CAFs while the increase in FAP^+^ CAFs was more pronounced, effectively lowering the αSMA/FAP ratio (**Fig. 3C-D, Extended Data Fig. 3A**). Immune profiling revealed an influx of both CD8^+^ and CD4^+^ T cells (**Fig. 3C, E, Extended Data Fig. 3B**), whereas Tregs are decreased in 5 out of 6 paired biopsies evaluated (**Fig. 3C, F, Extended Data Fig. 3C**).

**Figure 3.**
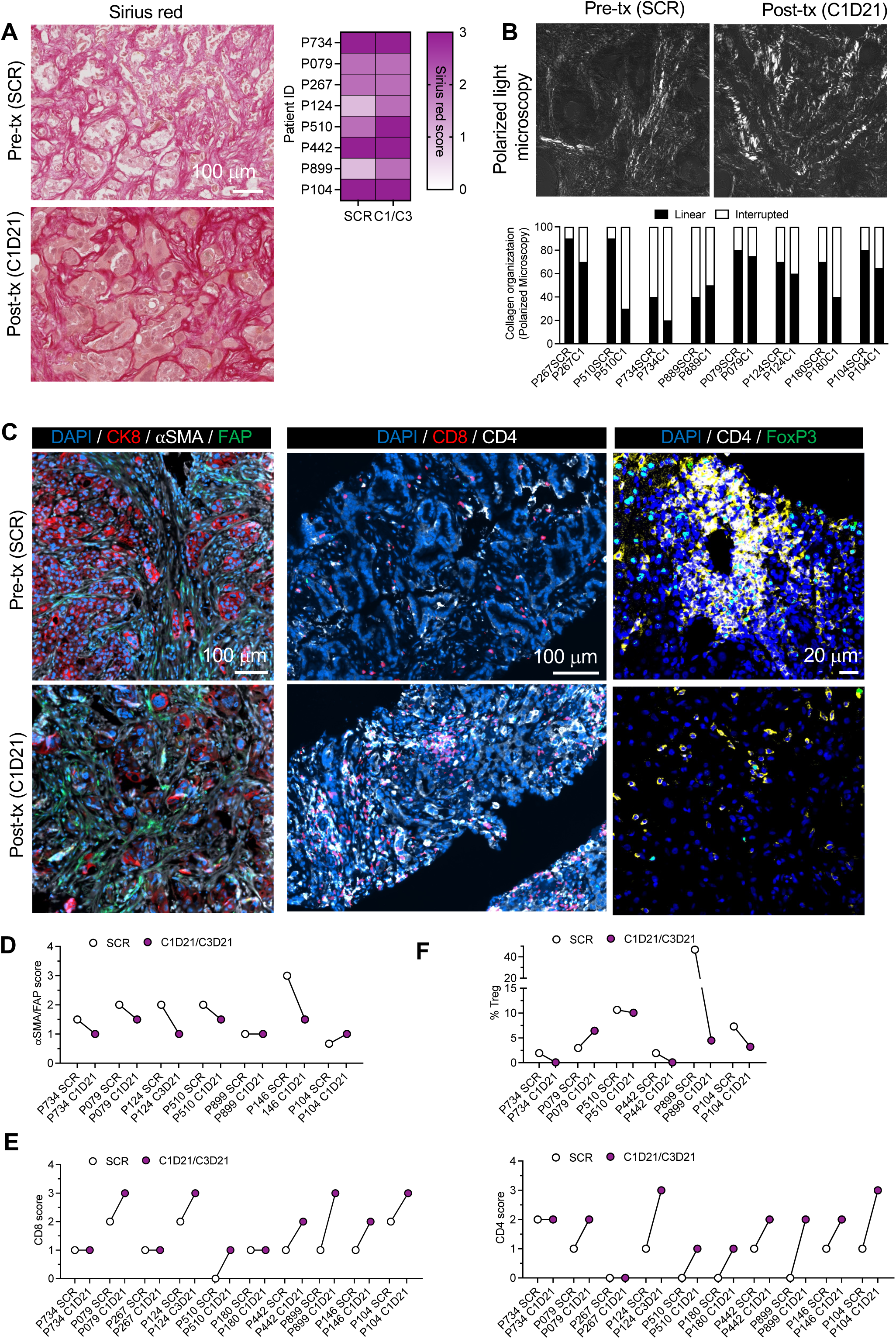
Hh priming remodels PDAC TME and increase intratumoral T cells. **A.** Representative sirius red stained paired biopsies pre-treatment (pre-tx, SCR = screening) and post-treatment (post-tx) biopsies and associated scoring. P124 consists of biopsy pairs at screening and C3D21; all others are screening and C1D21. **B.** Representative images of polarized light microscopy and associated quantification for linear vs interrupted collagen organization. **C.** Representative immunolabeling for the indicated markers for pre-treatment (pre-tx, SCR = screening) and post-treatment (post-tx) biopsies. **D.** αSMA/FAP ratio scores for the indicated paired biopsies. **E.** CD8^+^ T cells and CD4^+^ T cells scores for the indicated paired biopsies. **F.** FoxP3^+^ CD4^+^ Treg scores for the indicated paired biopsies.

Spatial transcriptomic analyses of selected biopsy pairs (**Fig. 4A**) from liver metastases or lymph nodes were carried out with delineation of cancer rich (IT: intratumoral), moderate (PM: peripheral margin) and low (DS: distant stromal) spots (**Extended Data Fig. 4A**). Post biopsies samples showed a consistent decrease in cancer cells compared to pre-treatment matched samples when selecting the cancer rich spots (IT), and with more post-treatment decrease noted in patient with greater antitumor response (P442, compared to P267, and then P510; **Fig 4B**, **Extended Data Fig. 4B**). The increase in FAP expressing CAFs was noted post-treatment (C1) compared to pre-treatment (screening, SCR), in particular in the peripheral margin (PM) (**Fig. 4B, Extended Data Fig. 4B**), consistent with immunolabeling studies (**Fig. 3C, D**). Also consistent with the immunolabeling studies (**Fig. 3C, D**), post-treatment samples showed minimal changes in Acta2 expressing CAFs compared to pre-treatment matched samples at the analyzed timepoint in the treatment scheme (αSMA CAF, **Fig. 4B**). Endothelial and perivascular cells enriched spots where heterogenous across matched samples pre- and post-treatment, though IT and PM spots showed a trend for increase in 2 of 3 paired biopsies (**Fig. 4B, Extended Data Fig. 4B**). An increase in memory CD4 effector T cells was consistently noted in IT spots post-treatment across all paired samples (**Fig. 4C, Extended Data Fig. 4B**), and –consistent with immunolabeling (**Fig. 3E**)– an increase in CD8^+^ T cells in IT, PM (**Fig. 4C**, **Extended Data Fig. 4B**), and DS spots. Exhausted CD8 T cells and Tregs show mixed responses across IT, PM and DS spots and biopsy pairs, though the biopsy pair from P442, captured from best responder in this spatial transcriptomic dataset (**Fig. 4A**), showed both Treg and exhausted CD8 T cells in IT spots (**Fig. 4C**, **Extended Data Fig. 4B**). Myeloid cells were increased post-treatment in IT and PM spots, whereas NK cells showed mixed composition pre- and post-treatment across spots and paired biopsies (**Fig. 4B**, **Extended Data Fig. 4B**). Gene expression analysis for tertiary lymphoid structure (TLS) showed enrichment post treatment in all paired biopsies (**Fig. 4D**). Differential gene expression post-treatment captured largely down regulation of gene expression, with pathway analyses indicating distinct pathway enrichment for each matched pair samples (**Extended Data Fig. 5**). P627 biopsy pair showed downregulation of genes associated with extracellular remodeling and ERK signaling, P442 showed more specific downregulation of genes associated with collagen and cellular cytoskeleton remodeling, and P510 showed downregulation in metabolic genes, in particular fatty acid and mitochondria metabolism (**Extended Data Fig. 5**). Taken together these results inform on the remodeling of the tumor microenvironment post-treatment favoring a milieu responsive to ICI, as well as enrichment of FAP expressing CAF. Notably heterogeneous transcriptomic changes are noted across paired biopsies following treatment, likely reflecting biopsy site sampling specific changes and heterogeneity in dynamic remodeling of the TME at a single sampling time point captured early during treatment.

**Figure 4.**
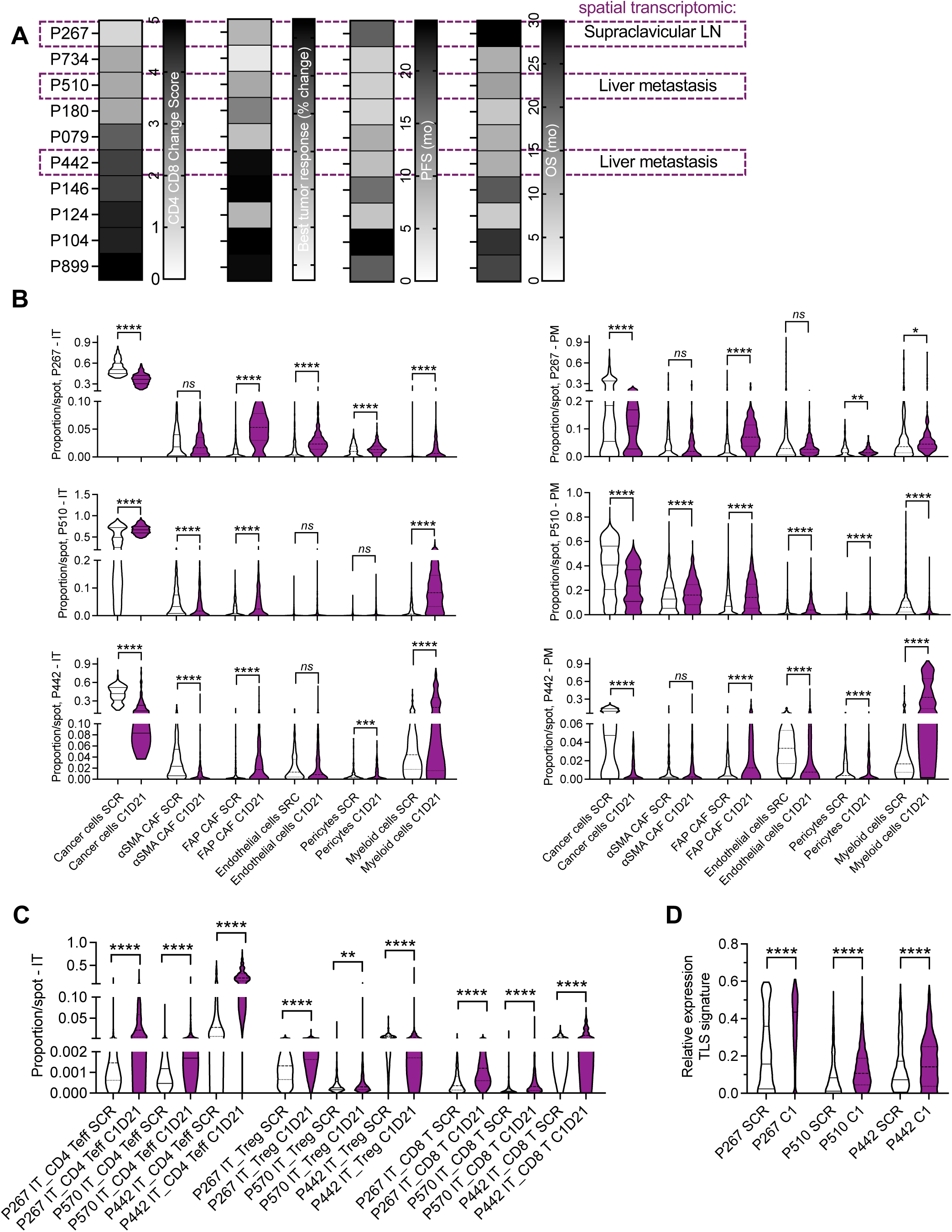
Spatial transcriptomics studies reveal polarized TME with Hh signaling targeting. **A.** Depiction of CD4 and CD8 score change in increasing order (higher score change depicts increased CD4 and CD8 T cell infiltration) and matched best percent tumor lesion size change, progression free survival (PFS) and overall survival (OS), highlighting the representation of paired samples selected for spatial transcriptomic analyses. Mo: months. **B-C.** Cell populations pre- and post-treatment in the intratumoral (IT) and peripheral margin (PM) spots for all paired biopsies. **D**. Tertiary Lymphoid structure (TLS) gene expression signature. Mann-Whitney test. * *P* < 0.05, **, *P* < 0.005, *** *P* < 0.001, **** *P* < 0.0001, *ns*: not significant.

## Discussion

Aberrant Hedgehog (Hh) signaling has long been implicated in pancreatic cancer biology, driving stromal activation and desmoplasia that foster immune evasion and therapeutic resistance^12–15,27,28^. Yet the net effect of Hh pathway modulation in PDAC is context dependent. Preclinical studies showed that Hh blockade can transiently enhance vascular perfusion and chemotherapy delivery, while simultaneously reshaping CAF states and, in some settings, promoting an immunosuppressive milieu^15,18–20,22,28^. Clinically, prior trials of Hh inhibitors (e.g., vismodegib, sonidegib, saridegib) failed to improve outcomes in PDAC, leading to discontinuation of development in this disease^15,28–30^. These mixed data argue that how and when the pathway is inhibited, particularly its timing relative to cytotoxic and immune therapies, may be decisive.

In this phase 1b/2 study, we operationalized an Hh ‘priming’ strategy with short, intermittent courses of the oral SMO inhibitor NLM-001 administered around cycles of gemcitabine/nab-paclitaxel together with the CTLA-4 inhibitor zalifrelimab. This design was informed by prior mechanistic work showing that depletion of αSMA⁺ CAFs or genetic loss of sonic Hh yielded less differentiated tumors and rendered the TME susceptible to checkpoint blockade^20,22^. Consistent with that premise, the combination in our trial produced encouraging clinical activity in the first-line metastatic setting (ORR 50%, mPFS 7.3 months), comparing favorably with historical benchmarks for GnP alone (ORR 23%, mPFS 5.5 months)^31^ and for GnP plus dual checkpoint blockade (ORR 30%, mPFS of 5.6 months)^32^, while acknowledging the limitations of cross-trial comparisons. Early molecular responses supported these findings with patients achieving on-treatment ctDNA clearance by cycle 4 experiencing longer PFS and OS, nominating serial ctDNA as a pragmatic pharmacodynamic biomarker for this regimen.

Correlative analyses suggest that transient Hh inhibition reshaped the PDAC stroma and immune contexture in a manner permissive to CTLA-4 blockade. Across paired biopsies, we observed collagen remodeling with less linear fiber architecture, a decrease in the αSMA/FAP ratio reflecting relative enrichment of FAP⁺ CAFs, and an influx of CD4⁺ and CD8⁺ T cells with reductions in Tregs. These histologic changes were mirrored by spatial transcriptomics showing decreased cancer cell relative abundance post-treatment and increased FAP-expressing CAFs with immune TME remodeling favoring expansion of memory CD4 and CD8 T cell populations in the intratumoral and peripheral margin. Gene expression analyses showed an increase in TLS signature post treatment, supportive of the predicted immunoresponsive TME post CAF priming treatment. Gene expression downregulation post treatment also informed on distinct pathways, although not uniformly across all sites, underscoring biologic heterogeneity by anatomic niche and timing. Taken together, these data are consistent with a model in which phasic Hh blockade transiently reduces contractility/ECM linearity and enhances a hypoxic and immunosuppressive TME, which – when coupled with chemotherapy and CTLA-4 inhibition – yields greater immune engagement. The heterogeneity we observed emphasizes that the CAF-immune axis in PDAC is dynamic and likely site-specific, and that optimal priming schedules may need to be tuned to maximize immune permissiveness while avoiding deleterious effects of stromal depletion.

Tolerability is a second point of differentiation. Historical experiences with Hh inhibitors, particularly when combined with chemotherapy, have been marked by substantial low-grade, cumulative toxicities that drive dose modifications and discontinuation^33–36^. In contrast, NLM-001 in our study was generally well tolerated, with predominantly grade 1–2 symptoms and no NLM-001 treatment-related discontinuations. While our trial was not designed to isolate the contribution of scheduling, the intermittent priming approach, together with the pharmacokinetic profile of NLM-001^25^, may have mitigated cumulative toxicity while preserving the desired stromal/immune effects.

This study has important limitations. It is a single-arm, early-phase trial with a modest sample size and investigator-assessed responses. The biopsies capture a single early on-treatment time point and predominantly sample liver metastases; both factors constrain generalizability of the TME findings. Additionally, although the efficacy signals exceed historical benchmarks, only a randomized comparison can determine incremental benefit over GnP (with or without CTLA-4 blockade) and define the independent contribution of NLM-001. Finally, while ctDNA clearance correlated with improved outcomes here, prospective validation is required before clinical adoption as a response-adaptation tool.

In conclusion, these data support a testable paradigm that precisely timed, phasic Hh inhibition can reprogram the PDAC stroma to enhance the efficacy of chemotherapy and CTLA-4 blockade, yielding meaningful responses with manageable toxicity. A randomized study with integrated spatial and liquid-biopsy biomarker analyses is warranted to confirm clinical benefit, refine the priming schedule, and identify patients most likely to respond to this stromal-immune co-targeting strategy.

## Methods

### Murine studies

*P48-Cre, LSL-Kras^G12D/+^, Tgfbr2^F/F^* (KTC) genetically engineered male and female mice (GEM) from age 35 to ∼65 weeks were housed in AAALAC-accredited facility at MD Anderson Cancer Center (MDACC), fed a LabDiet 5053 *ad libitum* and were maintained at 21-23°C, 40-60% humidity, and 12h light/dark cycle environment. Following genotyping and when mice reached 35 weeks of age, they were randomized to treatment with sonic hedgehog inhibitor (cyclopamine, LC Laboratories, 500mcL i.p. of at 40mg/kg in corn oil) vs diluent control (corn oil). Mice also received immune check point inhibitor (ICI) anti-CTLA 4 (BioXcell BE0131, clone 9H10, 200mcg i.p. for the first dose, then 100mcg i.p. for the second and third dose, every 3 days for 3 consecutive doses) and anti-PDL1 (BioXcell BE0101, clone 10F.9G2, 200mcg i.p., every 3 days for 3 consecutive doses) or control IgG (combination of BioXcell BE0090 clone LTF-2 and BE0087 polyclonal isotype control at matched dosing, respectively). Cyclopamine treated mice were also treated with ICI or IgG control and compared to corn oil treated mice. Mice were euthanized at the first sign of moribundancy. All procedures were reviewed and approved by the MD Anderson Cancer Center Institutional Animal Care and Use Committee (IACUC).

### Clinical trial

The Numantia trial is an open label, phase 1b/2, multicenter study that evaluates the efficacy, safety, and tolerability of NLM-001 in combination with gemcitabine and nab-paclitaxel (GnP) plus zalifrelimab (EudraCT: 2020-004932-52; NCT04827953). Patients with metastatic PDAC were enrolled and treated at six sites in Spain (**Supplementary Table 3**). Eligible patients had confirmed diagnosis of stage IV pancreatic adenocarcinoma (one patient had Stage III) that was previously untreated. Patients were aged 18 years or older with measurable disease per RECIST 1.1 and had an Eastern Cooperative Oncology Group (ECOG) performance status of 0 or 1 with adequate end-organ function. No prior treatment for metastatic disease was permitted. Patient 080 had surgical resection of ductal pancreatic adenocarcinoma and received FOLFIRINOX prior to enrolment into trial. Patients who have received chemotherapy for previously localized disease were eligible if at least six months have elapsed from the last chemotherapy treatment. The study was conducted in compliance with the Declaration of Helsinki and International Conference on Harmonization Guidelines for Good Clinical Practice and was approved by the institutional review board at each of the participating sites. All patients provided informed written consent.

Patients received conventional chemotherapy with gemcitabine (1000 mg/m^2^ IV) and nab-paclitaxel (125 mg/m^2^ IV) on days 1, 8 and 15 of each 28-day cycle according to standard of care. Zalifrelimab (Agenus AGEN1884) was given at a dose of 1mg/kg IV on day 15 of cycle 1 and every 6 weeks thereafter 30 min after administration of chemotherapy. NLM-001 was administered at the dose of 800 mg/day orally according to two schedules. In ‘Schedule 1’, NLM-001 was given for 4 days as a priming treatment before chemotherapy days 1 and 15 (on days - 4, - 3, - 2 and – 1, and on days 10, 11, 12, and 13), for cycles 1-3. In ‘Schedule 2” in which NLM-001 was given for three consecutive cycles followed by two rest cycles (cycles 1-3, 6-8, 11-13, 16-18 and so on). Paired biopsies were obtained, when feasible, for assessment of translational endpoints. Imaging was performed Q6W (±3 days) from the first treatment dose.

The primary end point was to assess objective response rate (ORR), defined as the proportion of patients experiencing complete response (CR) and partial response (PR) according to RECIST 1.1 based on investigator assessment. Secondary endpoints included the assessment of the following: safety and toxicity, according to the National Cancer Institute’s Common Terminology Criteria for Adverse Events (NCI-CTCAE) version 5.0; duration of response (DOR); disease control rate (DCR), defined as a complete response (CR), partial response (PR), or stable disease (SD) of ≥6 weeks; progression-free survival (PFS) and overall survival (OS), all per RECIST 1.1 based on investigator assessment.

### Treatment response evaluation by ctDNA-NGS monitoring

Tumor-informed ctDNA assay called TRACKseq (Altum Sequencing S.L.) was applied in plasma and whole blood cells collected at the time of diagnosis and after each cycle of treatment. Treatment response was assessed by monitoring ctDNA levels from same peripheral blood sample. First, the baseline mutational screening was performed on FFPE tumor biopsy samples using the Oncomine™ Colorectal and Pancreatic Panel. When solid biopsy was not available, a custom Ion Ampliseq HD panel was applied to plasma cfDNA for the molecular characterization of the tumor. All somatic and pathogenic genetic variants detected at any of the diagnosis samples analyzed were selected as tumor biomarker for both ctDNA and CTCs monitoring. An average of 2 somatic SNVs or INDELS were selected. A patient-specific multiplex PCR was designed including all suitable biomarkers and was applied to both fractions following a previously described protocol^37,38^. Libraries were sequenced on the Ion S5 System platform (Life Technologies, Thermo Fisher Scientific Inc.) and bioinformatically analyzed by a proprietary software. Same pipeline and mPCR was applied to three healthy control donor DNA samples to obtain the limit of detection (LOD) and limit of quantification (LOQ) for each biomarker experimentally. Samples with at least one variant detected were defined as ctDNA-positive. ctDNA status was defined by the mutation with the highest variant allelic fraction (VAF).

### Histopathology

Five-micrometer formalin-fixed, paraffin-embedded (FFPE) needle biopsies were deparaffinized and rehydrated. Hematoxylin and eosin (H&E) and Masson trichrome stain (MTS) were performed using staining kit (Leica Biosystems, 38016SS2). Nuclei staining was performed with Weigert’s hematoxylin followed by Sirius Red staining using Pico-sirius Red Solution (Direct Red 80, Sigma Aldrich 365548). All staining were performed according to the manufacturer’s direction. For murine histopathological analyses, H&E-stained tumor tissues were scored by delineating the relative percentage of normal (N), well-differentiated (W), moderate (M), and poorly (P) differentiated tumor. Representative images were captured using a Leica DM 1000 LED microscope equipped with a DFC295 microscope camera (Leica). The score was performed in a blinded fashion by a single operator. For SR morphometric scoring of human tissues, the slides were scanned using the Zeiss Axio Scan.Z1 430038-9001-000 and visualized using the ZEN 3.1 blue edition software. Focused scoring on intratumoral SR-stained area was performed using a score of 1 for 0-30% SR staining, 2 for 30-60%, and 3 for 60% or greater using 100x field of view. For polarized microscopy, Z1 fluorescence microscope was used as previously described^22^. Five random 200x images in the metastatic tumor, excluding normal liver parenchyma and portal triads, were analyzed using the NIH Image J software and data expressed as the relative percentage of linear vs. interrupted collagen organization.

### Immunolabeling and quantification

To analyze αSMA, CK8, and FAP (fibroblast panel); and CD4, FoxP3, and CD8 (immune panel), tyramide signal amplification (TSA) multiplex staining was performed. Five-micrometer formalin-fixed, paraffin-embedded (FFPE) needle biopsies were deparaffinized and rehydrated. Antigen retrieval was performed in TE buffer (pH 9.0) for 15 min at 98°C in a microwave. A hydrophobic barrier was then created around the tissue sections, followed by blocking with 4% CWFG for the vasculature panel and 1% BSA for the fibroblast and immune panels for 10 min at room temperature (RT). Blocking was followed by incubation with αSMA antibody (Dako 0651, 1:200) for the fibroblast panel, or CD4 antibody (Thermo MS-1528, 1:20) for the immune panel, each for 1 h at RT. Slides were then incubated with rabbit-on-rodent HRP-Polymer (Biocare Medical) for the vasculature panel and mouse-on-mouse HRP-Polymer (Biocare Medical) for the fibroblast and immune panels, for 10 min at RT. This was followed by staining with Opal 690 (Akoya Biosciences) for the fibroblast and immune panels, each for 10 min at RT. A second round of antigen retrieval was performed by heating slides in TE buffer (pH 9.0) at 98°C for 15 min in a microwave. Blocking was repeated as described above. Slides were then incubated with CK8 antibody (Millipore Sigma MABT329, 1:50) for the fibroblast panel, or FoxP3 antibody (Abcam Ab20034, 1:50) for the immune panel, overnight at 4°C. The following day, slides were incubated with rabbit-on-rodent HRP-Polymer for the vasculature panel and mouse-on-mouse HRP-Polymer for the fibroblast and immune panels, each for 10 min at RT. This was followed by staining with Opal 520 for the immune panel, for 10 min at RT. A third round of antigen retrieval was performed in TE buffer (pH 9.0) at 98°C for 15 min in a microwave. Blocking was repeated as previously described. Slides were then incubated FAP antibody (Abcam ab207178, 1:500) for the fibroblast panel, or CD8 antibody (Dako M7103, 1:100) for the immune panel, for 1h at RT. After primary antibody incubation, slides were treated with mouse-on-mouse HRP-Polymer for the fibroblast and immune panels, each for 10 min at RT. This was followed by staining with Opal 520 for the fibroblast panel, and Opal 570 for the immune panel, each for 10 min at RT. Finally, slides were incubated with Hoechst 33342 (Invitrogen, 1:10,000) for 10 min and mounted with Vectashield (Vector Laboratories). Images were acquired using the Zeiss Axio Scan.Z1 430038-9001-000 and the ZEN 3.1 blue edition software. For fibroblast panel, quantification was performed by scanning the entire section and assigning a score for intratumoral αSMA and FAP relative content on 50x field of view from a scale of 1 to 3, with a score of 1 assigned for 20% or less αSMA^+^ cells or 10% or less FAP^+^ cells, a score of 2 assigned for 20-50% αSMA^+^ cells or 10-30% FAP^+^ cells, and a score of 3 assigned for 50% or more αSMA^+^ cells or 30% or more FAP^+^ cells. A similar approach was used to score CD4^+^ and CD8^+^ intratumoral and directly peritumoral T cells (100x field of view) with a scale of 0 to 3, and a score of 0 assigned for 5 cells or less seen, a score of 1 assigned for 5-30 cells, a score of 2 assigned for 30-80 cells, a score of 3 assigned for 80 cells or more seen. Paired samples (SCR and C1/C3 biopsies) were excluded if one of the pair was missing tissue section to analyze, contained too little tumor tissue, or if staining was of poor quality preventing accurate scoring. FoxP3^+^ CD4^+^ relative quantification was performed by using the Zeiss LSM800 confocal microscope. The number of FoxP3^+^ CD4^+^ cells per up to nine field of views is expressed as a percentage of all CD4^+^ cells. The αSMA/FAP score was defined by dividing the αSMA score by the FAP score. The CD8^+^ T cell and CD4^+^ T cell change score was defined by dividing the C1/C3 CD8^+^ T or CD4^+^ T cell score by the SCR CD8^+^ T or CD4^+^ T cell score. If the SCR score is 0, the non-0 score of C1-C3 is kept. There were no ‘0’ score for C1/C3 samples alone for each pair. When both C1/C3 and SCR scores are ‘0’ for a pair, the score ratio is assigned a ‘0’ value. The sum of the CD8^+^ T cell and CD4^+^ T cell change score defines the ‘CD4 CD8 change score’.

### Spatial cell transcriptomics analyses

FFPE tissue blocks were scored and sectioned at 10 micron thickness. Deparaffinization and H&E staining was performed according to 10x Genomics protocol for Visium CytAssist Spatial Gene Expression for FFPE sample preparation (CG000518). H&E slides were scanned with a Zeiss AxioScan.Z1 and 11 x 11 mm regions for analysis were identified. Libraries were constructed using the Visium CytAssist Spatial Gene Expression for FFPE human kit (10X Genomics), pooled, and sequenced with the NextSeq500 (Illumina) per manufacturer instructions. Raw FASTQ files were aligned to the hg38 reference genome and spatially mapped using Space Ranger 2.0.1 (10x Genomics). For generating spatial cell type maps, we used Cell2location (version 0.1.3)^39^, which integrates Visium spatial data with scRNA-seq references. Cell2location applies a Bayesian framework to estimate the abundance of reference cell types at specific spatial coordinates, providing a decomposition of multi-cell spatial transcriptomics data. The analysis proceeded in several stages. First, a reference cell annotation was constructed from scRNA-seq data of human pancreatic liver metastases^40^, which was used as input to Cell2location. In the second step, gene selection was performed, and expression signatures were determined for each cell type. Subsequently, we used the default negative binomial regression model in Cell2location to estimate the reference cell-type signatures and predict cell type distributions at spatial locations. For the analysis of each Visium section, default parameters were applied, with the following exceptions: ‘max_spochs’ set to 100,000; and ‘N_cells_per_location’ set to 20. In addition, Cell2location was used to estimate the gene expression of each cell type at each spot by implementing an approach based on Robust Cell Type Decomposition (RCTD)^41^. The intratumoral (IT), peritumoral (PM, peripheral margin), and distant stromal (DS) regions were defined according to their spatial proximity to cancer-rich areas termed as cancer seeds, identified based on the cancer cell proportion derived from the Cell2location model. Cancer seeds were defined as spots within the top 30% of cancer cell proportions (quantile ≥ 0.70). The IT region comprised spots with 0 hops to the nearest cancer seed (tumor core), while the PM region included spots within 1-2 hops of the nearest cancer seed (adjacent tissue). The DS region encompassed spots with ≥3 hops from the nearest cancer seed (distant stromal tissue). Cell type proportions estimated by Cell2location were normalized to sum to 1 for each spot. The data is presented as violin plots generated in GraphPad prism. Differential gene expression analysis was conducted to identify genes with significantly altered expression between pre-and post-treatment samples. Gene expression data for cancer cells in both conditions were log-transformed (log1p) to address distribution skewness. A linear model was applied to each gene to assess changes in expression associated with the treatment. P-values were adjusted for multiple testing using the Benjamini-Hochberg method. The final differential expression statistics were compiled and used for subsequent pathway analysis using WebGestalt 2024^42^. For the TLS analysis, the Meylan signature^43^ was used in a gene set enrichment analysis (GSVA), with ssGSEA performed on each sample’s normalized expression data. Statistical comparisons between the SCR and C1 group were conducted using the Mann-Whitney test, and the results were summarized by patient and condition. The data was deposited into GEO, accession GSE312056, reviewers’ link:

https://urldefense.com/v3/__https://www.ncbi.nlm.nih.gov/geo/query/acc.cgi?acc=GSE312056__;!!PfbeBCCAmug!myrWzqUi7JCmlQrV3GqkxZfi8cLh_0snVKYJNNXEotzMDQSQ6AYElApqh1-2urnOdB2Cku_s5q9AXWjXqIUvJ9pfEBczDN5N$

### Statistical Analysis

All subjects who have received at least the first dose of the first cycle of treatment will be included in the safety analysis set (SAS). The efficacy evaluable set included all treated patients who received at least two complete cycles of study treatment. In this phase 1b/2 study, no formal sample size calculation was conducted. Enrollment of 28 subjects was considered reasonable to evaluate preliminary efficacy and safety of study combination. Descriptive statistics were used in general to summarize trial results, including means, medians, ranges, and measures of variability. PFS and OS were ascertained with Kaplan-Meier and Log-rank test using SPSS software version 26, or Gehan-Breslow-Wilcoxon test using GraphPad Prism 10. Median time to event(s) and 95% CI for the median are provided. The severity of AEs graded using NCI-CTCAE whenever possible. Graphs were generated using GraphPad Prism and statistical analyses used are listed in the respective figure legend.

## Data Availability

De-identified clinical data from individual participants that support the findings presented in this study are available for academic use upon request, subject to the constraints of the informed consent provided. Researchers interested in accessing these data can submit a request, along with a signed data transfer agreement, to the corresponding author (Manuel Hidalgo). Access will be granted to those who agree to the terms outlined in the data transfer agreement, which include using the data solely for research purposes and ensuring the confidentiality of patient and data information for the duration of the agreement. The data will be accessible for requests up to 2 years following the conclusion of the Numantia study. Source data for each figure is provided in the Source Data File. https://urldefense.com/v3/__https://www.ncbi.nlm.nih.gov/geo/query/acc.cgi?acc=GSE312056__;!!PfbeBCCAmug!myrWzqUi7JCmlQrV3GqkxZfi8cLh_0snVKYJNNXEotzMDQSQ6AYElApqh1-2urnOdB2Cku_s5q9AXWjXqIUvJ9pfEBczDN5N$

## Acknowledgements

The authors are grateful to the patients who participated in this study and their families. The authors also thank APICES for the project management, monitoring, data management and statistics assistance. The authors thank Julienne L. Carstens and Kathleen M. McAndrews for assistance with murine and human tissue initial assessment, respectively.

## Data Sharing Statement

De-identified clinical data from individual participants that support the findings presented in this study are available for academic use upon request, subject to the constraints of the informed consent provided. Researchers interested in accessing these data can submit a request, along with a signed data transfer agreement, to the corresponding author (Manuel Hidalgo). Access will be granted to those who agree to the terms outlined in the data transfer agreement, which include using the data solely for research purposes and ensuring the confidentiality of patient and data information for the duration of the agreement. The data will be accessible for requests up to 2 years following the conclusion of the Numantia study. Source data for each figure is provided in the Source Data File.

## Conflict of interest statement

In this study, the compound NLM-001 was supplied by Nelum Pharma. MH is a stock equity owner and serves as the Chairman of the Advisory Board for Nelum Pharma. MH also serves on the Board of Directors of Bristol Myers Squibb (BMS) and Guardant Health (GH). MH also declares stock and other ownership interests in Champions Oncology and InxMed, receives honoraria from Champions Oncology and Velavigo, has consulting or advisory role on OncoMatrix and InxMed, leadership roles at BMS and Guardant Health, receives funding from TopAI, Ranok Therapeutics, Incyte, and receives paid expenses from BMS and Guardant Health and royalties from Myriad for PALB2 patent. EP is the whole stock equity owner in I-xploremyhealth Corp. I-xploremyhealth is an equity owner in Nelum Corp. EP serves as the Chief Executive Officer and Chairman of Board for Nelum Pharma. EP is also an equity owner in PH Research, SL. EP serves as the Chief Executive Officer and Chairman of Board for Nelum Corp. VSK and RK are stock equity holders in PranaX, Inc. VSK and RK are founders, stock owners, and scientific consultants to PranaX, Inc. BB reports Research Funding from Agenus Inc and NanoView Bioscience; Travel Expenses from Erytech Pharma and Agenus; Advisory Board and Consulting from SpringWorks, Blueprint Medicines, BioLine Rx, Boxer and Enlivex; Speaker bureau from Total Health. The remaining authors declare no competing interests.

---

**Extended Data Figure 1.**
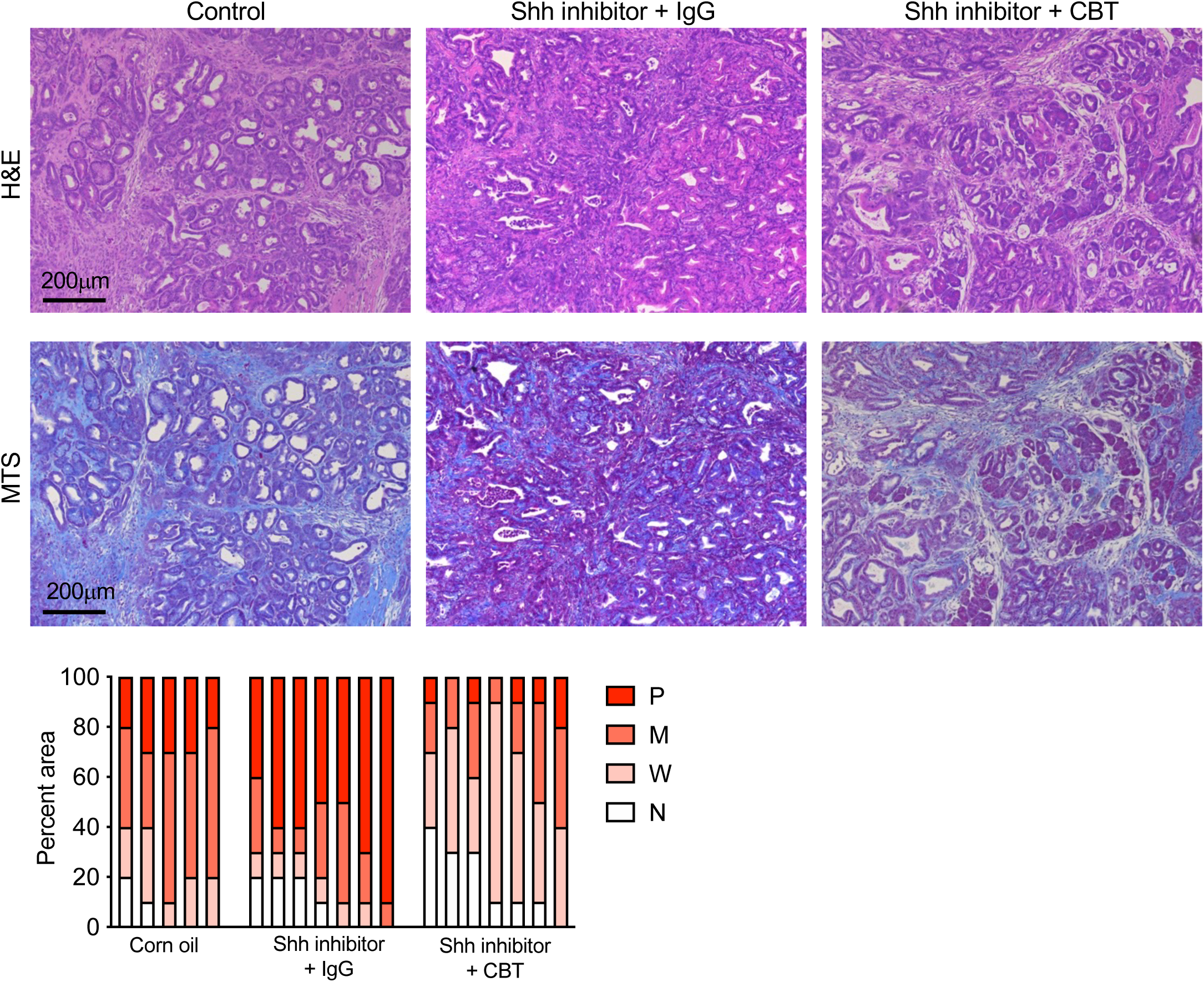
Hh signaling targeting with cyclopamine enables ICI response. Representative H&E and MTS stained PDAC tumor from mice in the indicated treatment group. Shh inhibitor: cyclopamine. CBT: anti-CTLA-4/anti-PD1 combination immune check point inhibitor (ICI). IgG: ICI control. Bar graph depicting relative area of the tumor section with poorly (P), moderate (M), well (W), and normal (N) histopathological features.

**Extended Data Figure 2.**
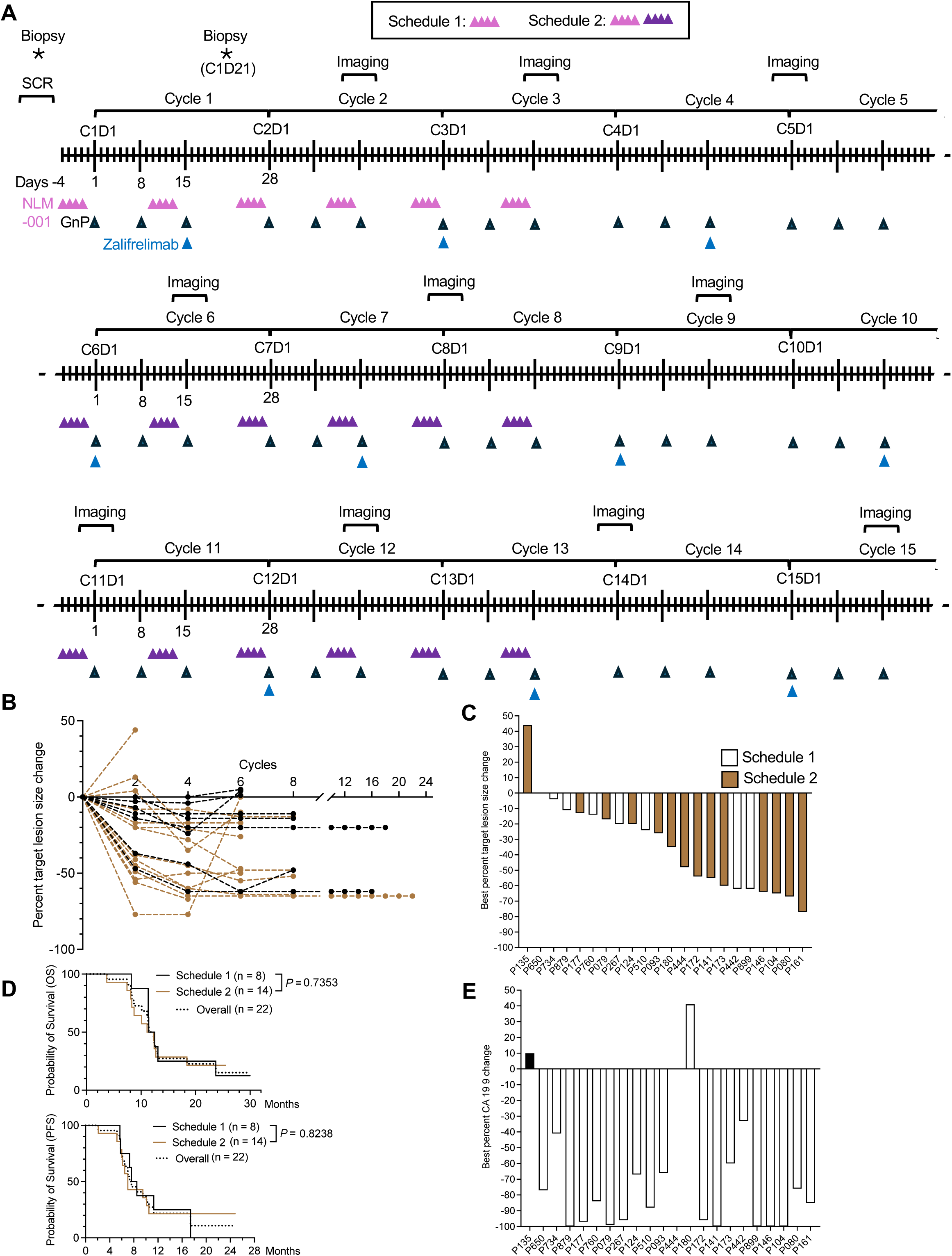
Detailed clinical trial treatment schedules. **A.** Detailed cycle definition for the listed therapies, repeated NLM-001 priming cycles in schedule 2 (C1-C3, C6-C8, C11-C13, C16-C18 and so on) vs a single priming in schedule 1 (C1-C3). **B**. Percent change in target lesions ascertained overtime for each patient on study (black: schedule 1; brown: schedule 2). **C**. Bar graph depicting best percent target lesion size change for each patient with defined allocation to schedule 1 vs schedule 2. **D.** Progression free survival (PFS) and overall survival (OS) of patients allocation to schedule 1 vs schedule 2. Log-rank test. Mo: months. **E**. Bar graph depicting best percent change in CA 19-9 for each patient listed with patient distribution matching the order of best percent change in tumor lesion size (C).

**Extended Data Figure 3.**
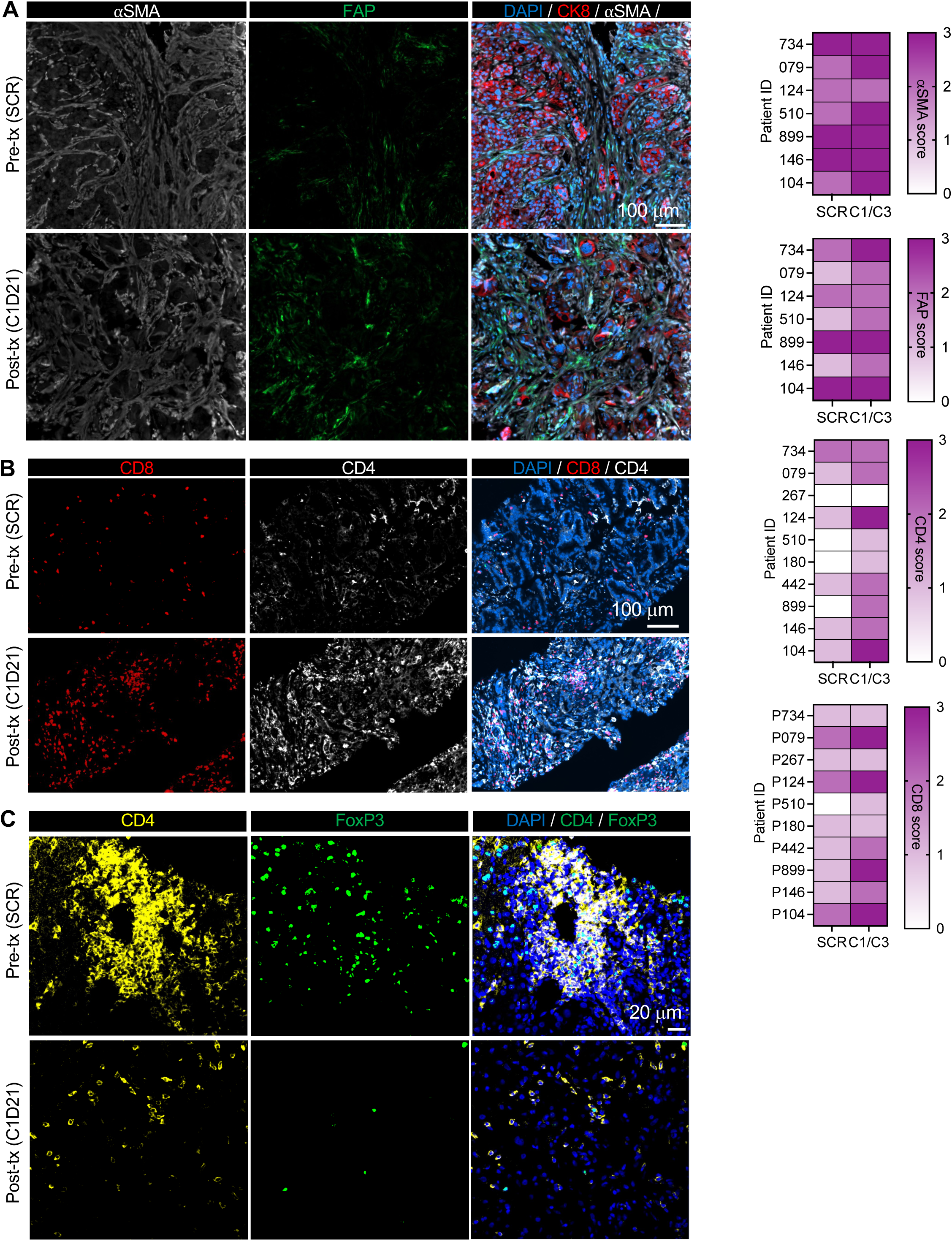
Immunolabeling of immune TME in paired biopsies. **A-C.** Representative immunolabeling for the indicated markers for pre-treatment (pre-tx, SCR = screening) and post-treatment (post-tx) biopsies from Fig. 3C with split channels, and depiction of scores from Fig. 3C as heat maps. *Related to Fig. 3*.

**Extended Data Figure 4.**
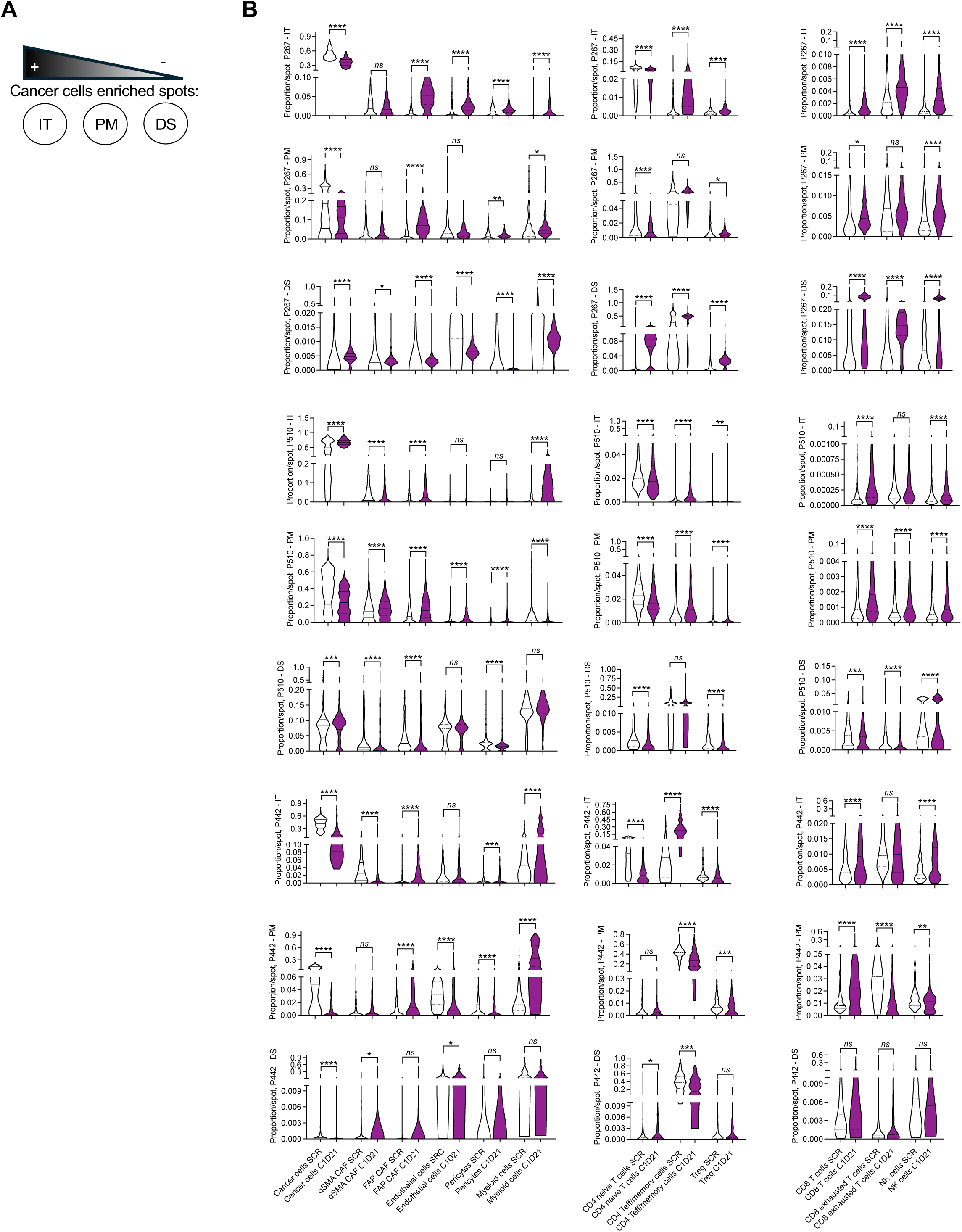
Spatial transcriptomic analyses of paired biopsies. **A.** Schematic representation of spatial transcriptomic spot with differential relative enrichment of cancer cells and defined as intratumoral (IT), peripheral margin (PM), or distant stromal (DS) regions. **B.** Cell populations pre- and post-treatment for all paired biopsies in each spot. Mann-Whitney test. * *P* < 0.05, **, *P* < 0.005, *** *P* < 0.001, **** *P* < 0.0001, *ns*: not significant. *Related to Fig. 4*.

**Extended Data Figure 5.**
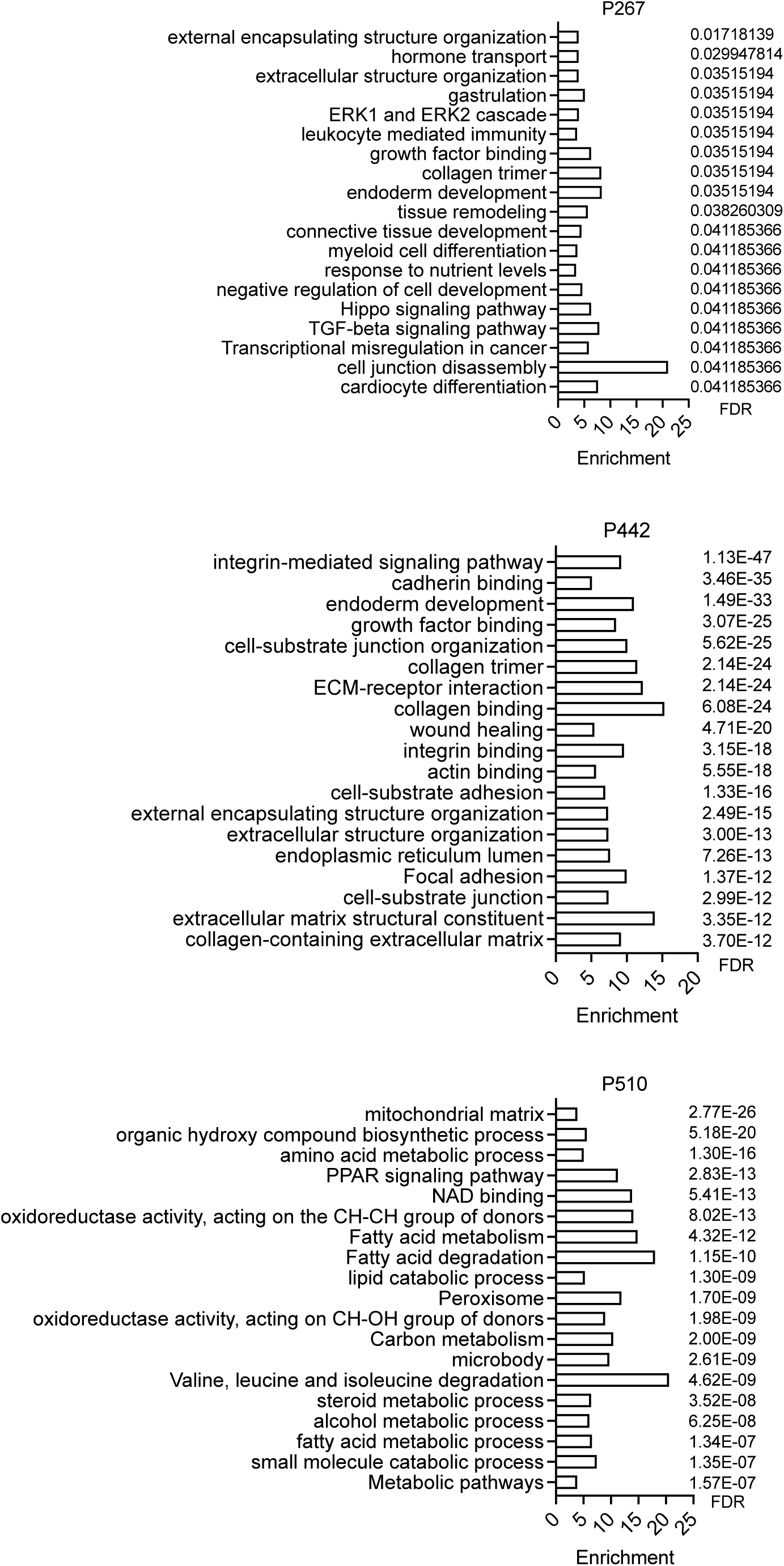
Gene expression pathways in spatial transcriptomic analysis of paired biopsies. Gene expression pathways for downregulated genes comparing pre- and post-treatment paired biopsies. Enrichment informs on number of genes in each pathway, FDR: false discovery rate.

**Extended Data Table 1.** Paired biopsies information.

| Extended Data Table 1. Paired biopsies information |  |  |  |  |  |  |  |  |  |
| --- | --- | --- | --- | --- | --- | --- | --- | --- | --- |
| Patient ID | Timepoint |  |  | Scheme |  | Clinical Response |  |  |  |
|  | Screening | C1D21 | C3D21 | Scheme 1 | Scheme 2 | PFS (mo) | OS (mo) | Survival after progression (mo) | Tumor Response |
| P079 | ✓ | ✓ |  |  | ✓ | 9.6 | 11.1 | 1.5 | SD |
| P104 | ✓ | ✓ |  |  | ✓ | 24 | 27.6 | 3.6 | PR |
| P124 | ✓ |  | ✓ |  | ✓ | 7 | 7.4 | 0.4 | SD |
| P146 | ✓ | ✓ |  |  | ✓ | 15.8 | 25.2 | 9.4 | PR |
| P173 | ✓ | ✓ |  |  | ✓ | 6.3 | 12.3 | 6 | PR |
| P180 | ✓ | ✓ |  |  | ✓ | 5.2 | 8.1 | 2.9 | PR |
| P267 | ✓ | ✓ |  | ✓ |  | 17.3 | 30 | 12.7 | SD |
| P442 | ✓ | ✓ |  | ✓ |  | 7.4 | 11.3 | 4 | PR |
| P510 | ✓ | ✓ |  | ✓ |  | 5.8 | 13.1 | 7.2 | SD |
| P734 | ✓ | ✓ |  | ✓ |  | 5.7 | 11.4 | 5.7 | SD |
| P899 | ✓ | ✓ |  | ✓ |  | 17.4 | 23.7 | 6.2 | PR |
| SD: stable disease; PR: partial response |  |  |  |  |  |  |  |  |  |
| mo: month |  |  |  |  |  |  |  |  |  |

**Extended Data Table 2.** Treatment Allocation.

| Extended Data Table 2. Treatment allocation |  |  |  |  |  |  |  |
| --- | --- | --- | --- | --- | --- | --- | --- |
|  |  | Scheme 1 (N=12) |  | Scheme 2 (N=16) |  | Total (N=28) |  |
|  |  | N | % | N | % | N | % |
| NLM-001 administered | Yes | 12 | 100.0 | 16 | 100.0 | 28 | 100.0 |
|  | Total | 12 | 100.0 | 16 | 100.0 | 28 | 100.0 |
| Zalifrelimab administered | No | 2 | 16.7 | 2 | 12.5 | 4 | 14.3 |
|  | Yes | 10 | 83.3 | 14 | 87.5 | 24 | 85.7 |
|  | Total | 12 | 100.0 | 16 | 100.0 | 28 | 100.0 |
| Nab-Paclitaxel administered | No | 1 | 8.3 | 0 | 0.0 | 1 | 3.6 |
|  | Yes | 11 | 91.7 | 16 | 100.0 | 27 | 96.4 |
|  | Total | 12 | 100.0 | 16 | 100.0 | 28 | 100.0 |
| Gemcitabine administered | No | 1 | 8.3 | 0 | 0.0 | 1 | 3.6 |
|  | Yes | 11 | 91.7 | 16 | 100.0 | 27 | 96.4 |
|  | Total | 12 | 100.0 | 16 | 100.0 | 28 | 100.0 |

**Extended Data Table 3.**
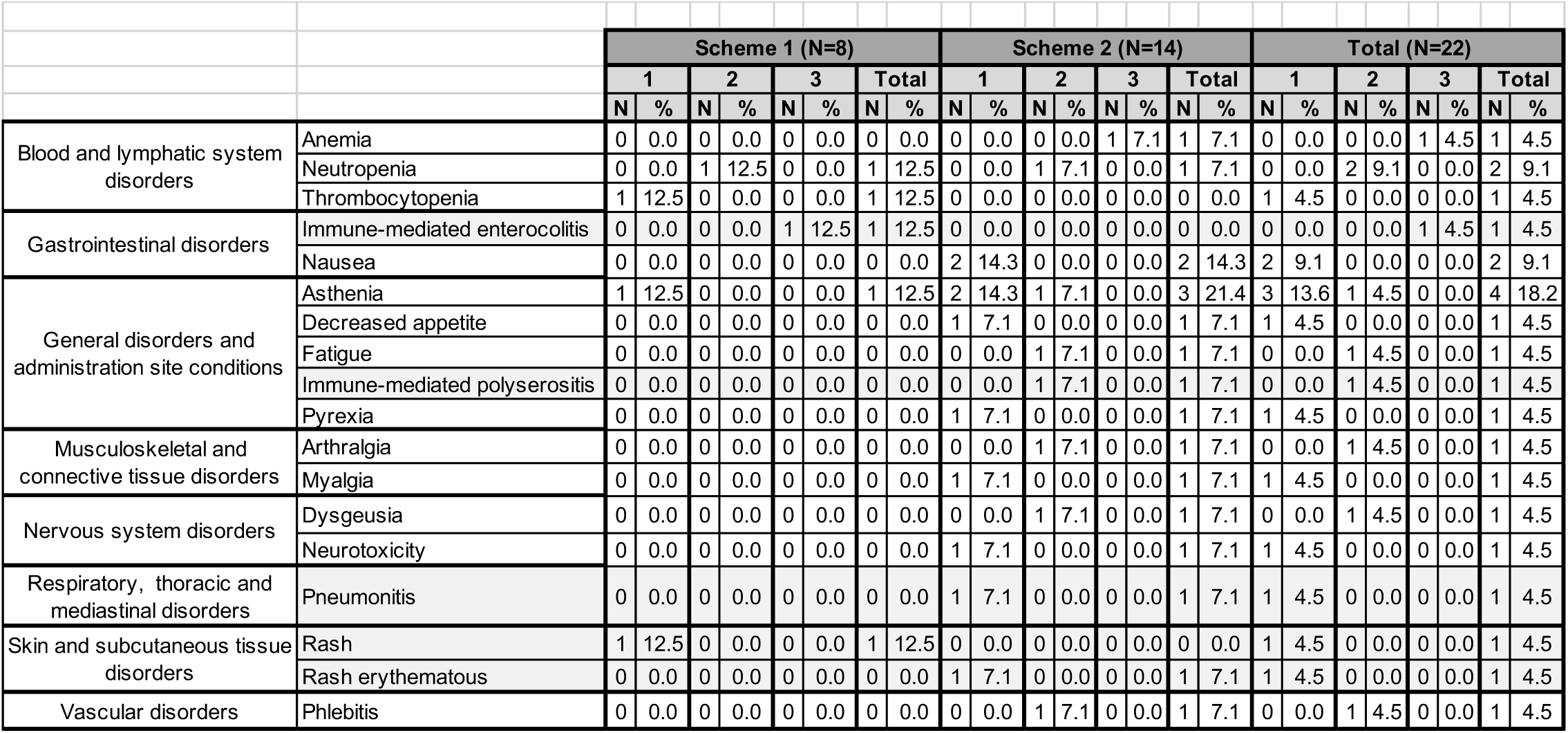
Adverse events related to zalifrelimab (alone or in combination with other study drugs) by max grade and scheme.

**Extended Data Table 4.**
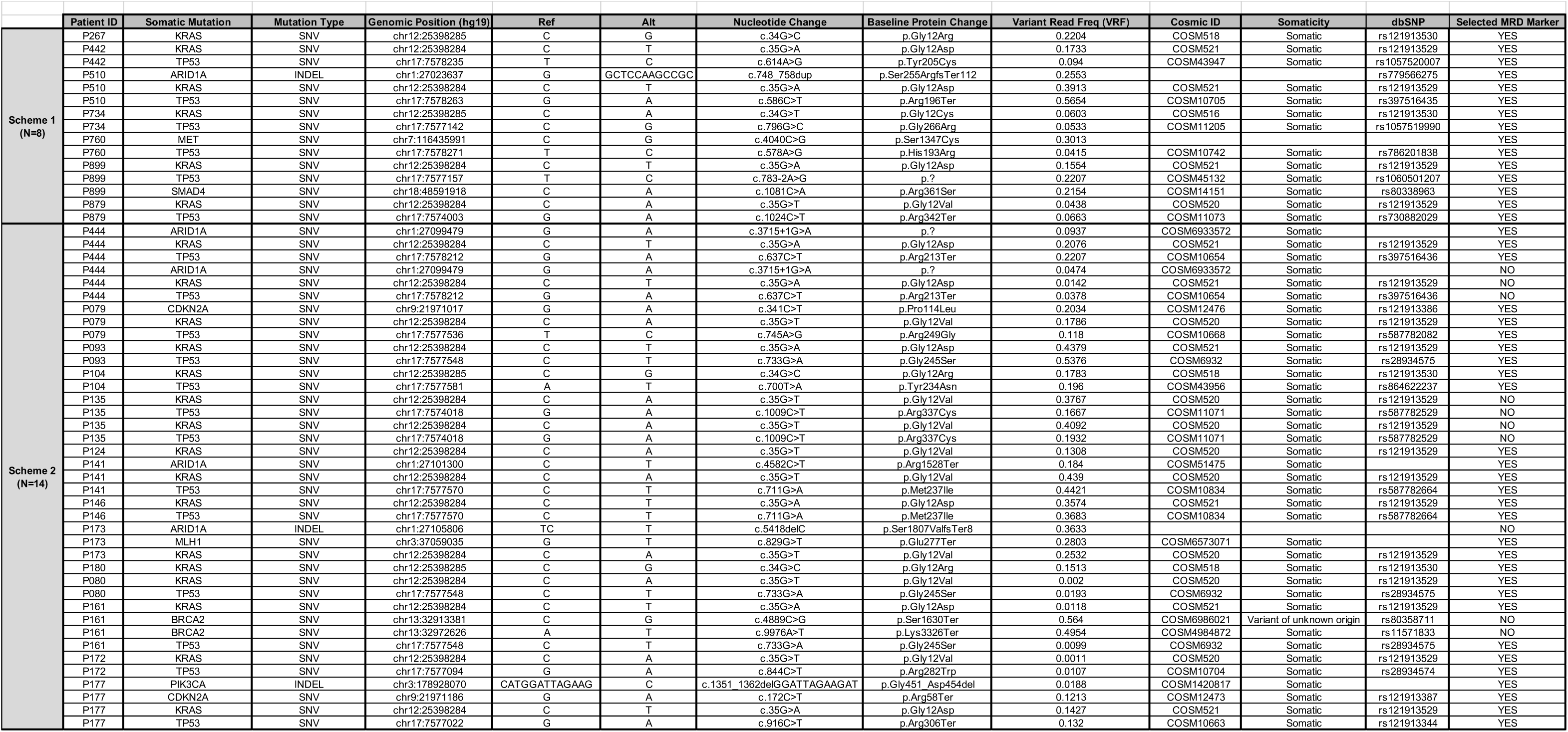
Mutations followed on ctDNA analysis.

**Supplementary Table 1.**
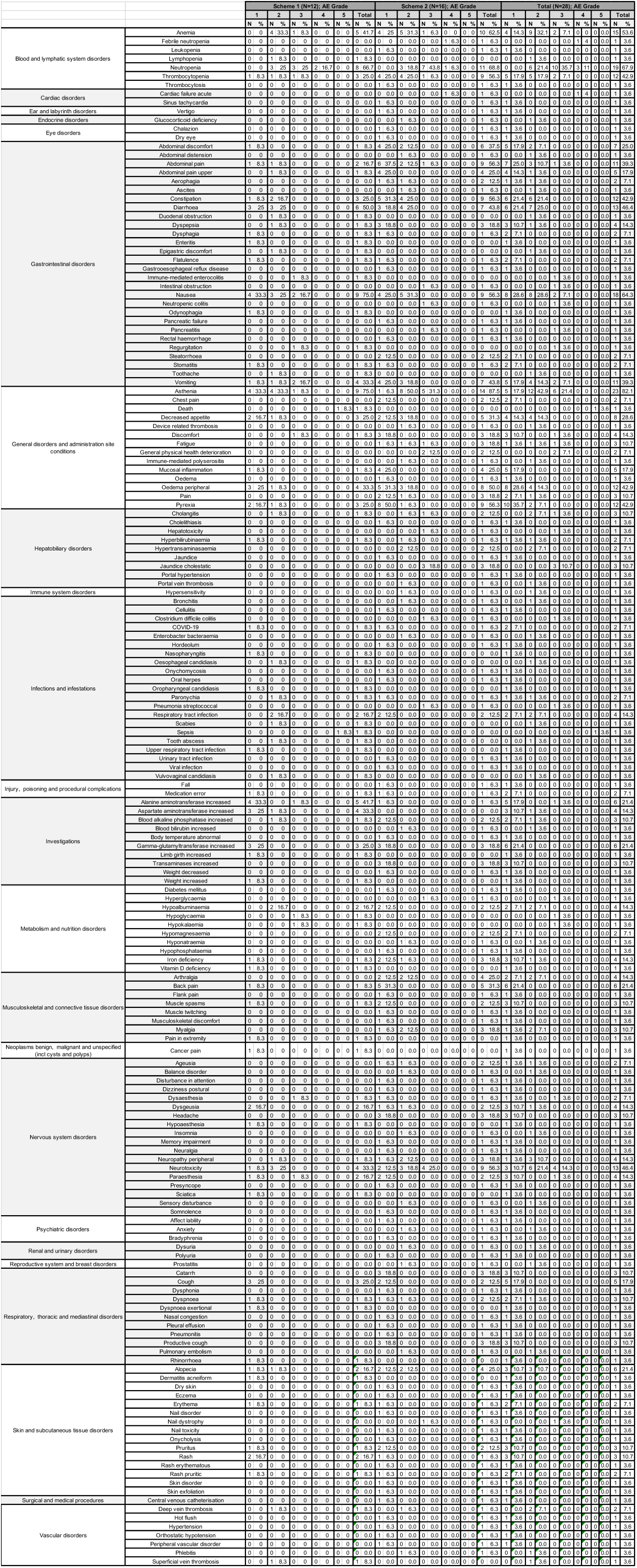
Treatment emergent adverse event (TEAE)

**Supplementary Table 2.**
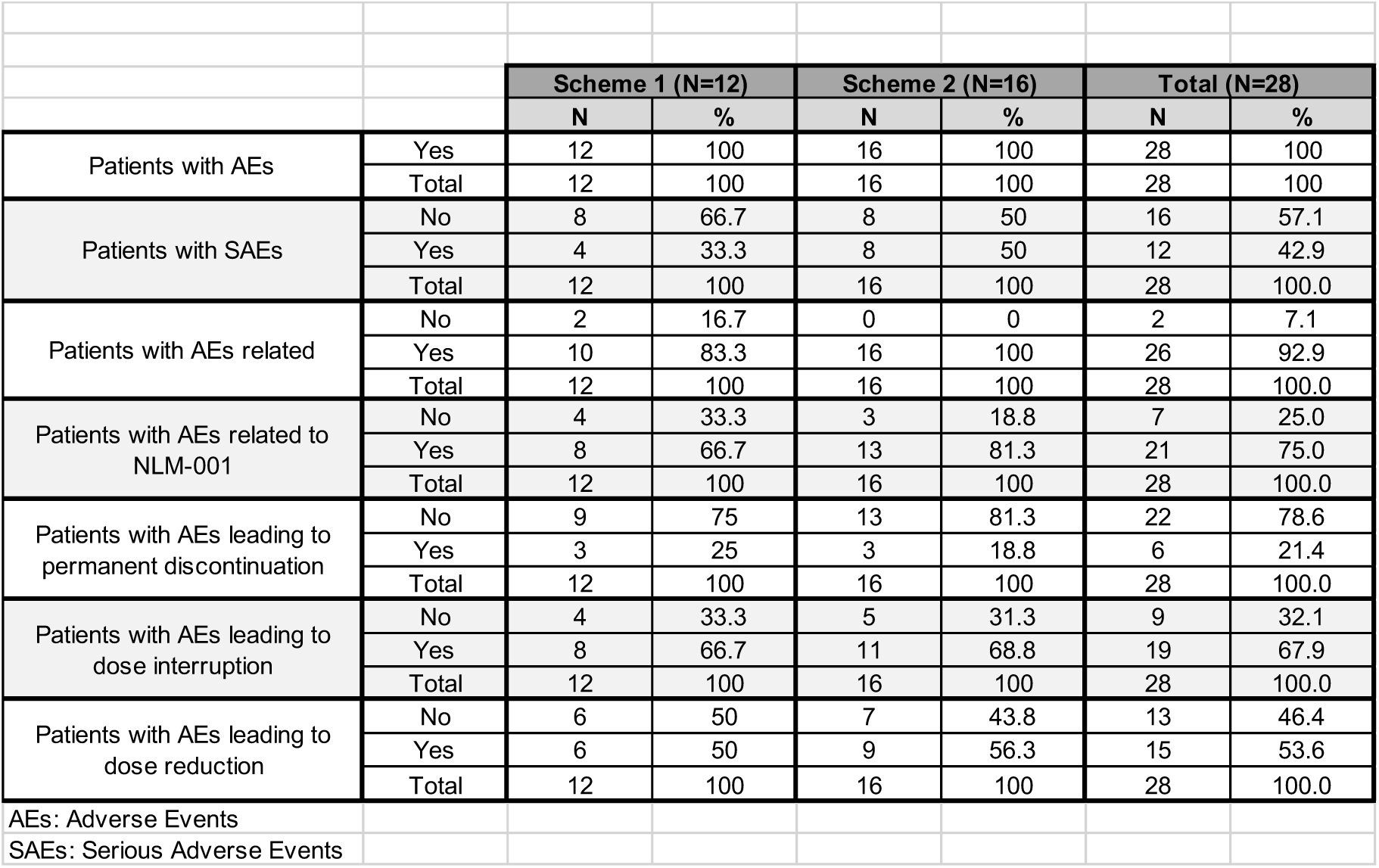
Adverse event (AE) encountered in at least one patient.

**Supplementary Table 3.** Clinical Trial Sites.

| Supplementary Table 3. Clinical Trial Sites |  |  |  |  |  |  |  |
| --- | --- | --- | --- | --- | --- | --- | --- |
| Safety Study (N = 28) | Site | N | % |  |  |  |  |
|  | HOSPITAL UNIVERSITARI VALL D'HEBRON | 5 | 17.9 |  |  |  |  |
|  | HOSPITAL VIRGEN DE LA VICTORIA | 6 | 21.4 |  |  |  |  |
|  | HOSPITAL UNIVERSITARIO MIGUEL SERVET | 7 | 25.0 |  |  |  |  |
|  | CENTRO ONCOLÓGICO DE DONOSTIA | 6 | 21.4 |  |  |  |  |
|  | CHU DE SANTIAGO | 2 | 7.1 |  |  |  |  |
|  | HOSPITAL UNIVERSITARIO MARQUÉS DE VALDECILLA | 2 | 7.1 |  |  |  |  |
|  | Total | 28 | 100 |  |  |  |  |
|  |  | Scheme 1 (N=8) |  | Scheme 2 (N=14) |  | Total (N=22) |  |
| Efficacy Study (N = 22) | Site | N | % | N | % | N | % |
|  | HOSPITAL UNIVERSITARI VALL D'HEBRON | 2 | 25.0 | 2 | 14.3 | 4 | 18.2 |
|  | HOSPITAL VIRGEN DE LA VICTORIA | 4 | 50.0 | 1 | 7.1 | 5 | 22.7 |
|  | HOSPITAL UNIVERSITARIO MIGUEL SERVET | 1 | 12.5 | 5 | 35.7 | 6 | 27.3 |
|  | CENTRO ONCOLÓGICO DE DONOSTIA | 1 | 12.5 | 3 | 21.4 | 4 | 18.2 |
|  | CHU DE SANTIAGO | 0 | 0.0 | 1 | 7.1 | 1 | 4.5 |
|  | HOSPITAL UNIVERSITARIO MARQUÉS DE VALDECILLA | 0 | 0.0 | 2 | 14.3 | 2 | 9.1 |
|  | Total | 8 | 100 | 14 | 100 | 22 | 100 |

## Notes

### Clinical Trial

NCT04827953

### Clinical Protocols

https://clinicaltrials.gov/study/NCT04827953

### Funding Statement

The study did not receive any funding

### Author Declarations

The study was conducted in compliance with the Declaration of Helsinki and International Conference on Harmonization Guidelines for Good Clinical Practice and was approved by the institutional review board at each of the participating sites. All patients provided informed written consent. The participating institutions are: HOSPITAL UNIVERSITARI VALL D'HEBRON HOSPITAL VIRGEN DE LA VICTORIA HOSPITAL UNIVERSITARIO MIGUEL SERVET CENTRO ONCOLOGICO DE DONOSTIA CHU DE SANTIAGO HOSPITAL UNIVERSITARIO MARQUES DE VALDECILLA

